# Wireless Colorimetric Multi-Biomarker Sensing to Enable Critical Neonatal Monitoring

**DOI:** 10.1101/2025.09.21.25336258

**Authors:** Alejandra Castelblanco, Elisabetta Ruggeri, Giusy Matzeu, Motaharehsadat Heydarian, Kai Foerster, Ahmed Bahnasy, Andreas Flemmer, Julia A. Schnabel, Benjamin Schubert, Fiorenzo G. Omenetto, Anne Hilgendorff

## Abstract

Clinical monitoring in the most vulnerable patients such as newborns relies on invasive and costly procedures and/or wired sensor surveillance, increasing discomfort and risk for undetected events. Addressing this critical need, we present a noninvasive, biocompatible silk-based sensor capturing multiple critical body functions via colorimetric analysis of body fluids, optimized for its use in critically ill preterm neonates where transepidermal fluid mirrors the interstitial compartment, thereby opening new avenues for future clinical monitoring. The sensor introduces twelve different colorimetric inks that stabilize labile bioresponsive molecules to a simple, wax-printed paper microfluidic wearable patch to facilitate real-time tracking of biomarkers (temperature (T), pH, sodium (S), and glucose (G)) on a miniaturized surface. Colorimetric responses showed high precision and reproducibility in sensing ranges relevant for neonatal care (T: 32-41 [°C]; pH: 3-9; S: 2.92-29.20 [mg/mL], G: 0.039-0.625 [mg/mL]). Deep learning for color response quantification achieved automated measurement (T:0.455 [°C]; pH:0.416; S:0.857 [mg/mL], G:0.019 [mg/mL]; mean absolute error). The sensor’s optimized interface for sample collection on skin and its performance under clinically relevant conditions, e.g., increased humidity (80%), tracking on moving object (0.986 AP @ IoU=0.5), sensor shear/rotation, uncontrolled light, as well as successful capture of physiological effects in biological fluids together with its miniaturized design caters to the critical needs of intensified monitoring in high-end patient populations such as neonates.

**Significance Statement:** We designed a noninvasive, wearable and miniaturized paper sensor patch capturing critical body functions via colorimetric analysis of body fluids (perspiration and saliva) adapted to the clinical needs of neonatal monitoring. Through twelve different bioresponsive inks stabilized in silk-fibroin into a simple, wax-printed paper microfluidic patch, the sensor facilitates simultaneous real-time tracking of multiple biomarkers under clinically relevant conditions using deep learning models for automated sensor detection and parameter estimation.

Graphical Abstract

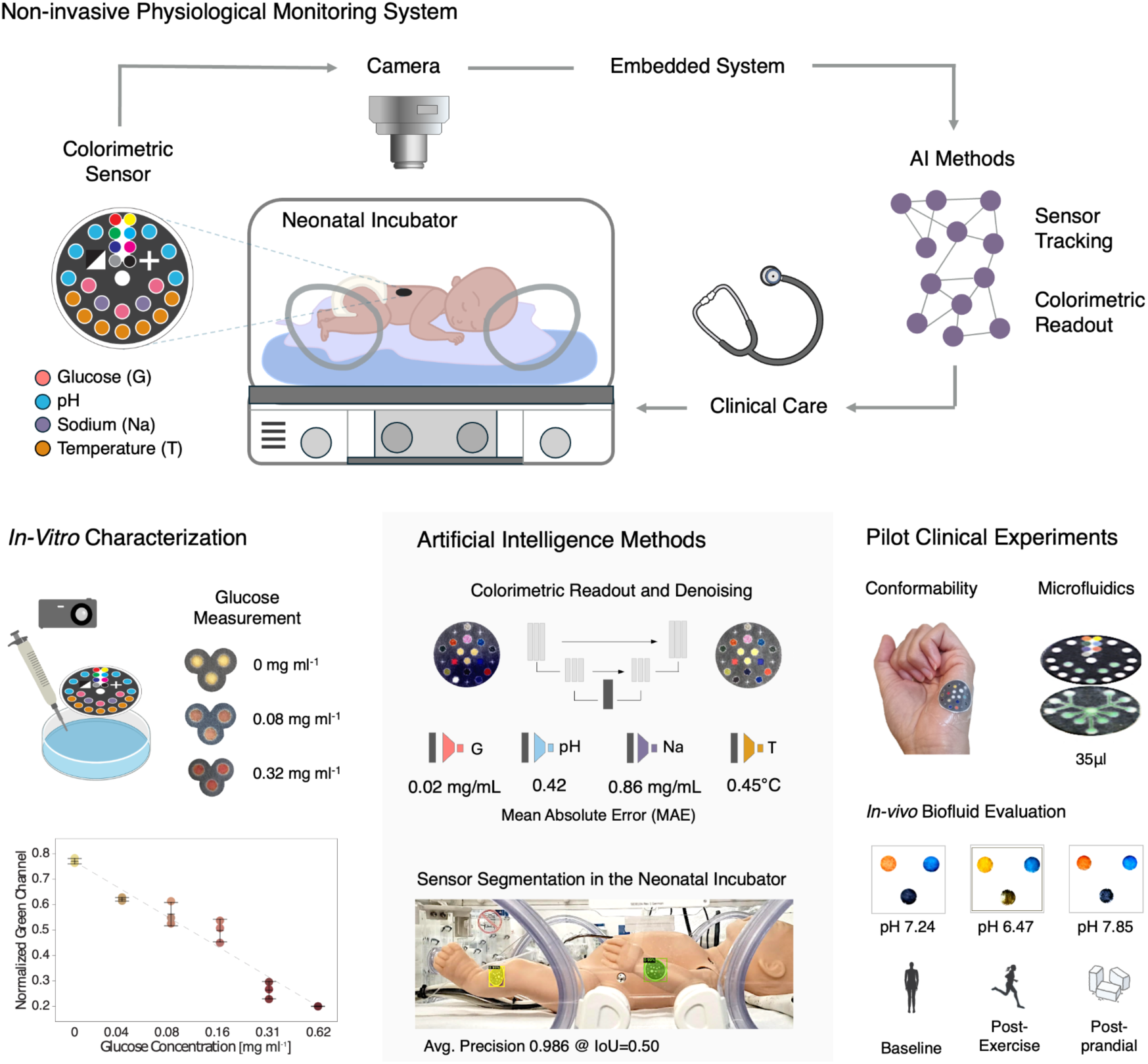

## 1. Introduction

Monitoring critical functions of the human body with easy-to-use, wireless biofluid sensors that are carefully adapted to the critical needs of specific patient populations will revolutionize patient care as their application reduces the need for invasive procedures, extensive laboratory equipment, and costly production. The most challenging showcase for the design of such devices is their application to neonates after at-risk birth, as they require parallel monitoring of different body function biomarkers through miniaturized devices, ensuring skin conformability, biocompatibility, and clinical usability while reducing production costs or troublesome maintenance.

As of today, however, neonatal care depends on a comprehensive set of medical devices and laboratory tests to assess vital signs and metabolic (e.g., glucose/electrolyte homeostasis) as well as organ function (e.g., liver, kidney, heart) for the prevention of (undetected) complications and timely treatment decisions. Devices for non-traumatic, continuous monitoring in these patients are only available for distinct parameters such as blood oxygenation and temperature [1]-[2]. These devices are wire-dependent and exclude assessment of other relevant metabolic functions, thus increasing the risk for undetected electrolyte and glucose imbalances [3]-[6], as well as increasing the delayed diagnosis of organ dysfunction (e.g., cystic fibrosis, liver failure) [6]-[7]. Not only is the use of wired sensors in neonatal intensive care known to obstruct daily care tasks and reduce parental skin bonding [8], but the subsequent need for invasive procedures also results in limitations for health care and trauma for the newborn and its family. Blood extractions are limited by sample volume and number as they require painful vessel punctures, increase the risk of critical blood loss, infection, and employ costly laboratory analysis [1], [2], [6], [9].

Easily accessible body fluids such as perspiration and saliva remain largely unexplored for clinical, noninvasive health monitoring in pediatric patients [9]. One of the few examples is iontophoresis, where perspiration fluid is used for diagnostic purposes in infants with cystic fibrosis. Although successfully implemented in routine care, chemical sample analysis still requires laboratory equipment and holds the risk of skin irritation due to electric stimulation [9]. Likewise, continuous neonatal glucose monitoring in interstitial fluid, via subcutaneous biosensors, has shown improved glucose level control in clinical studies, with remaining risks of subcutaneous inflammation, needle insertion pain, sensor drift, and recalibration requirements [10]. Making biological fluids even more valuable for diagnostic strategies, transdermal fluid in the preterm neonate in part reflects the interstitial and circulatory compartment [11], as skin immaturity increases insensible water loss, reaching up to permeation rates of 15-22 g/m^2^/hr [12], [13]. In adults, *in situ* perspiration analysis through potentiometry or amperometry has been explored, although it is limited by the reliance of these tools on energy harvesting modules, low device flexibility, higher weight, and significant maintenance restricting their application in routine clinical care [14], [19].

The possibility to advance clinical care by the use of high-end inventions in the bioengineering field was only taken up in a few devices that explored the use of microfluidic structures for passive biological fluid collection and chemical analysis [20], [21], including polymeric or paper-based colorimetric sensors [22]-[26]. The use of flexible and absorbent materials is crucial to ensure effective fluid sampling while avoiding skin irritation through traction. Not yet meeting clinical needs especially in critical patient populations, previous studies have mainly focused on detecting single analytes, and only few studies have focused on colorimetric wearable multi-biomarker sensing [27]-[30], or longitudinal sampling [31]. No studies, however, have successfully adapted the sensor in function and design to the specific challenges of neonatal monitoring, thereby missing opportunities noninvasive, light weight paper-based colorimetric sensing devices can offer for critical patient populations.

Adding to the complexity of the clinical solution needed, current colorimetric sensors are often limited by relying on human visual interpretation, which, influenced by illumination conditions and the visual acuity of the examiner, can result in significant measurement errors [32], [33]. Therefore, artificial intelligence (AI)-based image analysis and robust colorimetric signal digitalization [34], [35] represent a promising alternative for standardized, objective, and automated neonatal monitoring in the clinic, thereby alleviating the workload of nurses and other healthcare professionals. Advanced AI models are needed to integrate challenges posed by uncontrolled light conditions in the clinical setting, as well as demonstrate efficacy for tracking the sensor on a moving patient, as minimal handling and free range of motion for the patient is desired in pediatrics and especially neonatal care.

To address the clinical need for noninvasive, wireless monitoring of health indicators in the challenging conditions brought forward in critically ill neonates, we developed miniaturized microfluidic silk-based colorimetric sensors and combined them with a deep-learning imaging system to enable the noninvasive measurement of multiple critical physiological parameters in perspiration fluid, accompanied by demonstrating its alternative use in saliva (***Fig. 1a***). For neonatal monitoring, we achieved measurements of biomarkers that reflect critical functions in four different categories: (1) temperature as an example for critical body function, (2) glucose for energy consumption, (3) pH for metabolic balance, and (4) sodium for fluid balance/electrolyte homeostasis in clinically relevant ranges. To improve sensing reliability, we tested different chromogenic compounds for each analyte. Furthermore, accurate digital colorimetric analysis was achieved with our deep-learning imaging pipeline, even in uncontrolled light conditions. Working towards direct clinical application, key aspects of clinical feasibility, including microfluidic performance, dye stability in the neonatal incubator, and experimental application with biological fluids were investigated, demonstrating the sensor’s potential for future clinical application.

**Figure 1.**
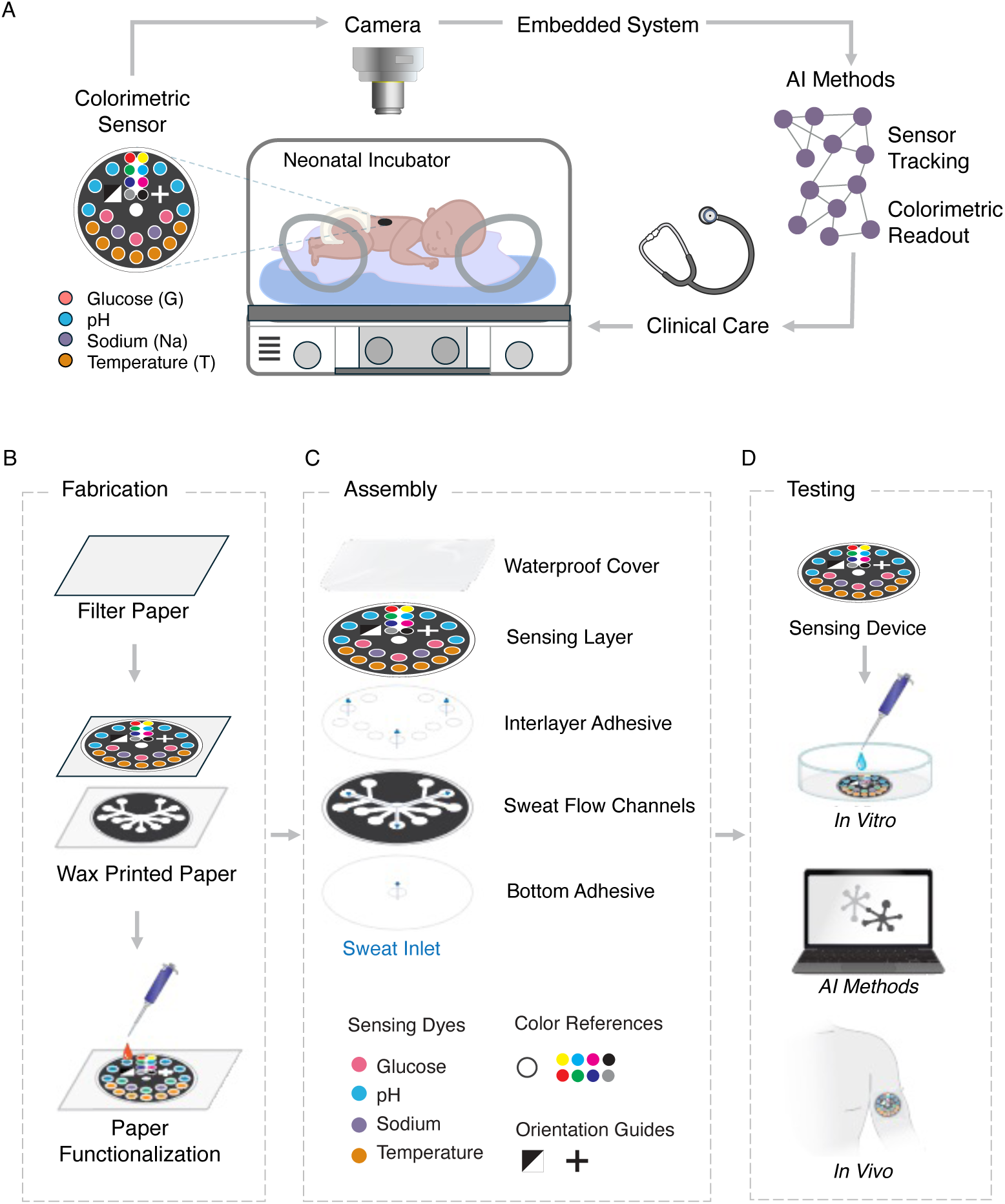
Neonatal non-invasive physiological monitoring system for perspiration biomarkers via colorimetric sensors. **A.** Schematic of the monitoring system including: a colorimetric paper-based sensor adhered to the newborn skin, a camera for image acquisition inside the incubator, an embedded system for image analysis, and a set of artificial intelligence (AI) models for sensor tracking and color-based parameter readout. **B.** Colorimetric sensor fabrication involves wax printing on a silk fibroin film and paper functionalization with colorimetric dyes. **C.** The sensor is assembled with three main layers: the sensing layer displaying the colorimetric dyes, the intermediate layer containing multiple fluid flow channels, and the bottom layer with a fluid inlet. On top, a waterproof medical skin cover film improves dye stability and skin adhesion. **D.** The sensor’s clinical feasibility is evaluated by characterization of the colorimetric response with *in vitro* experiments, AI-model testing for sensor tracking and parameter estimation, and experimental validation with *in vivo* biological fluids.

## 2. Results

Advanced bioengineering allowed multiple sensing areas in a redundant fashion for validity on a miniaturized sensor patch using a skin-contact optimized bottom sample inlet. *In vitro* evaluation of the colorimetric sensing array demonstrated accurate and reproducible measurement of temperature, pH, glucose, and sodium within physiologically relevant ranges for neonates. Automated parameter estimation was achieved using a deep-learning imaging model and tested in various illumination conditions. A pilot validation demonstrated adequate measurement of parameters with adult perspiration and saliva samples. Additionally, the functionality of the sensor was confirmed in simulated neonatal intensive care conditions, including sensor operation within a neonatal incubator.

### A. Colorimetric Sensing Technology and Experimental Characterization

The sensor patch was built using silk-fibroin as a biocompatible structural material (diameter of 2.8 cm; thickness of ∼0.2 mm; weight of 0.05g). The sensors integrate three main layers: the top layer displays the sensing dyes, the intermediate layer holds multiple fluid flow channels, and the bottom layer contains the fluid inlet (***Fig. 1b-c***). A transparent, water-resistant, and medically approved adhesive film covers the sensor, minimizing sample leakage and contamination. The sensor integrates a total of 12 different colorimetric dyes (7 temperature, 3 pH, 1 glucose, and 1 sodium) printed over individual circular areas for display of the color response (***Fig. 1b-c***). Dyes are made of the biocompatible materials silk and chitosan that are combined with enzymes (e.g., glucose oxidase and horseradish peroxidase) and pigments (e.g., pH, temperature indicators) to enable the colorimetric response to the biological target (*Materials and Methods, b*). In extensive *in vitro* experiments, the color response was recorded upon exposure to specific target concentrations in standard solutions or controlled temperature conditions, respectively using clinically relevant ranges for each parameter (***Table S1***). Color values were extracted through image analysis to generate calibration curves and to determine the dyes’ sensitivity and effective sensing range (*Materials and Methods, b)*.

For *temperature* measurements, the sensitivity, quantified as grayscale response [0–1] per °C, and the sensing range were measured for the seven colorimetric dyes (***Fig. 2a-c***): T1 (0.194, 33-36°C), T2 (0.140, 35-38°C), T3 (0.064, 32-41°C), T4 (0.070, 35.5-41°C), T5 (0.046, 36-41°C), T6 (0.137, 32-35.5°C, and 0.040, 36-41°C), T7 (0.032, 37-40°C), with the integrated response of all dyes enabling measurements in the range from 32°C to 41°C. The color response was reproducible in simultaneous measurements with three different sensors, the maximum standard deviation (SD) in the grayscale response [0–1] for each dye was: T1_SD_=0.068, T2_SD_=0.055, T3_SD_=0.023, T4_SD_=0.020, T5_SD_=0.012, T6_SD_=0.032, T7_SD_=0.014.

**Figure 2.**
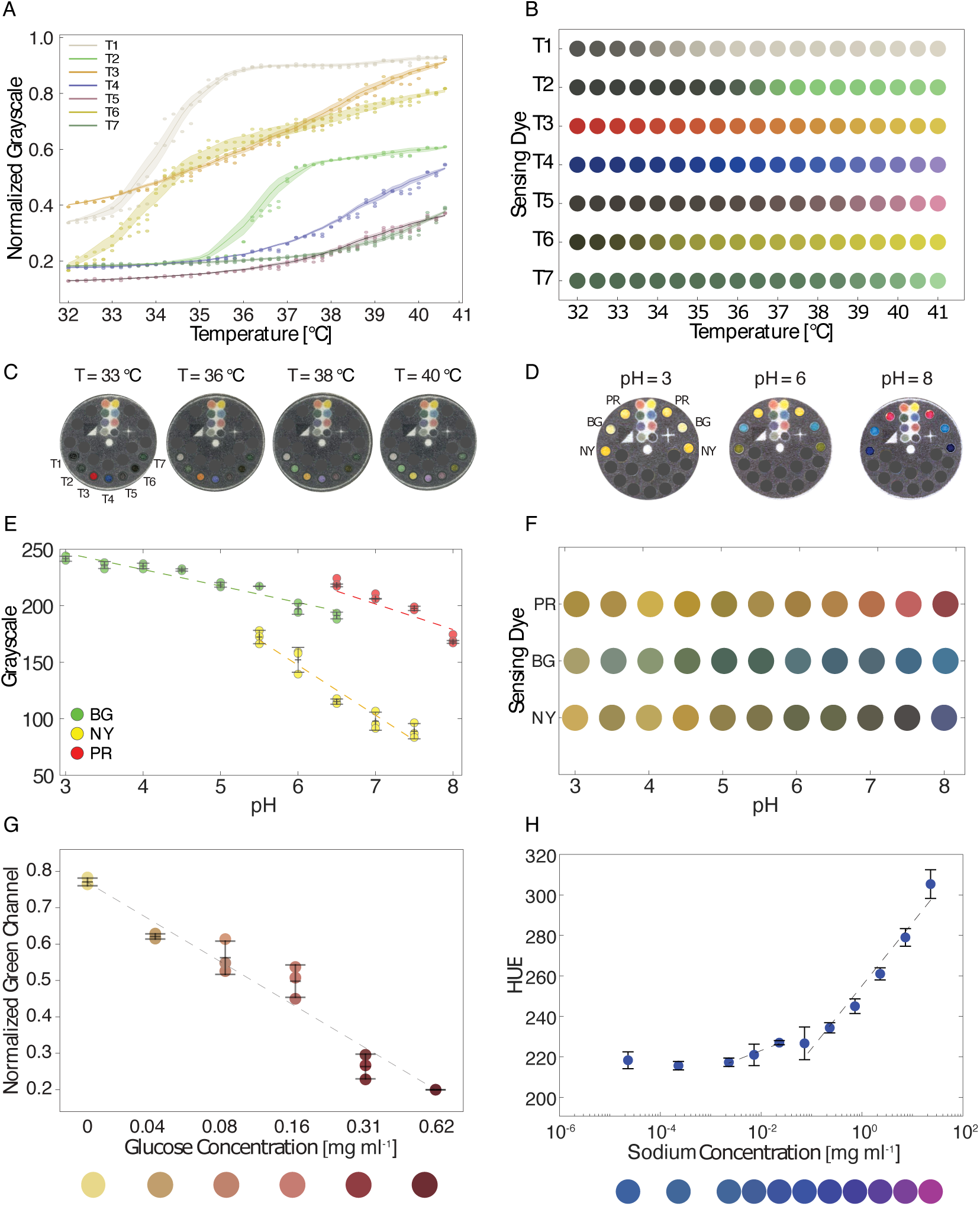
Colorimetric response for each analyte, including sensing range and sensitivity (normalized color/parameter units). **A.** Temperature Colorimetric Dye Response in the range 32 – 41 °C with sensitivities (norm. grayscale/°C) of: T1 (0.19), T2 (0.14), T3 (0.06), T4 (0.07), T5 (0.05), T6 (0.14), T7 (0.03). **B.** Temperature color scale for colorimetric temperature-responsive dyes (T1-T7). **C.** Experimental colorimetric responses for temperature at 33°C, 36°C, 38°C, 40°C. **D.** Experimental colorimetric responses for pH at 3, 6, 8. **E.** pH Colorimetric Dye Response in the range 3-8 with sensitivities (grayscale) of: PR (−0.07), BG (−0.09), NY (−0.12). **F.** pH color scale for three colorimetric temperature-responsive dyes (PR, BG, NY). **G.** Glucose Colorimetric Dye Response in the range 0 – 0.62 mg/mL with sensitivity of –1.95 (norm. green channel/mgmL^-1^). **H.** Sodium Colorimetric Dye Response in the range 2.90 – 29.0 mg/mL with sensitivity of 2.81 (hue/mgmL^-1^).

Colorimetric measurements of *pH* were performed via three different sensing dyes (PR, BG, NY), each exhibiting a unique colorimetric response (***Fig. 2d-f***), with the respective sensitivities (normalized (norm.) grayscale/pH) and pH sensing ranges of: PR (−0.073, 4.5-9), BG (−0.094, 3-5.5) and NY(−0.117, 4-7), demonstrating viability within the relevant pH range for clinical applications.

Characterization of the *glucose* colorimetric dye showed a sensitivity of –1.95 (norm. green-channel/mgmL^-1^) with a sensing range from 0.039 mgmL^-1^ to 0.625 mgmL^-1^, while the SD of the green-channel intensity for three replicates was between 0.013 to 0.081 (***Fig. 2g***). The *sodium* colorimetric dye displayed a sensitivity of 0.014 (norm. red-channel/mgmL^-1^) and 2.81 (hue/mgmL^-1^) in the sensing range of 2.92 mgmL^-1^(0.05 molL^-1^ NaCl) to 29.20 mgmL^-1^(0.5 molL^-1^ of NaCl), with a SD of the red-channel intensity between 0.031 to 0.075 (***Fig. 2h***).

The temperature colorimetric response was examined by exposing the sensors to continuous temperature cyclic variations in the measurement range. All the temperature dyes (T1-T6) showed cyclic, i.e., bidirectional reversible color responses (***Sup. Fig. 2a***), and hysteresis was bound below 0.025 in the normalized grayscale [0–1], equivalent to 0.78°C for the T3 dye.

For the pH dyes (PR, BG, NY) cyclic color changes were observed in a laboratory set-up with continuous exposure to a changing pH solution (*Materials and Methods, b*), grayscale [0–1] hysteresis for the pH dyes was PR (0.065 ± 0.026), BG (0.103 ± 0.064) and NY (0.112 ± 0.055; average ± SD), equivalent to 0.55 pH units for the PR dye (***Sup. Fig. 2b***).

For the pH and sodium inks, reversibility depends on their binding equilibrium, and further evaluation in complex environments is required [36]. In contrast, glucose-responsive dyes enable measurement by undergoing a binding-induced signal change that reaches saturation.

Unlike many conventional detection agents, silk-based materials are chemically stable and do not undergo non-specific redox reactions under regular operating conditions. Therefore, long-term storage stability and retained colorimetric response in pH, temperature, and glucose have been previously proven for silk-fibroin supported dyes for 24 months under refrigerated conditions (4°C), and 2.5 months for pH and temperature in accelerated aging conditions (60°C) [25], [36], and at least 8 days for glucose at (25°C) [37].

### B. Automated AI-based Colorimetric Measurement

A deep-learning model based on the U-Net [38] architecture was developed for automated estimation of the colorimetric sensor response, accounting for realistic conditions such as variable perspectives, sensor shear deformation, rotation, and non-uniform illumination across the sensor surface. The U-Net-based module denoises the image and corrects illumination artefacts, while a multi-layer perceptron (MLP) module is jointly trained to do regression of the target parameter values based on the corrected latent representation of the sensor (***Fig. 3a***), thus enabling simultaneous illumination correction and value estimation in 0.151 ± 0.015 seconds per image (CPU Apple M1, 16GB RAM). As a baseline to compare performance for color-based parameter estimation; a simpler convolutional neural network (CNN) combined with an MLP was trained and evaluated per variable (*Materials and Methods, c*).

**Figure 3.**
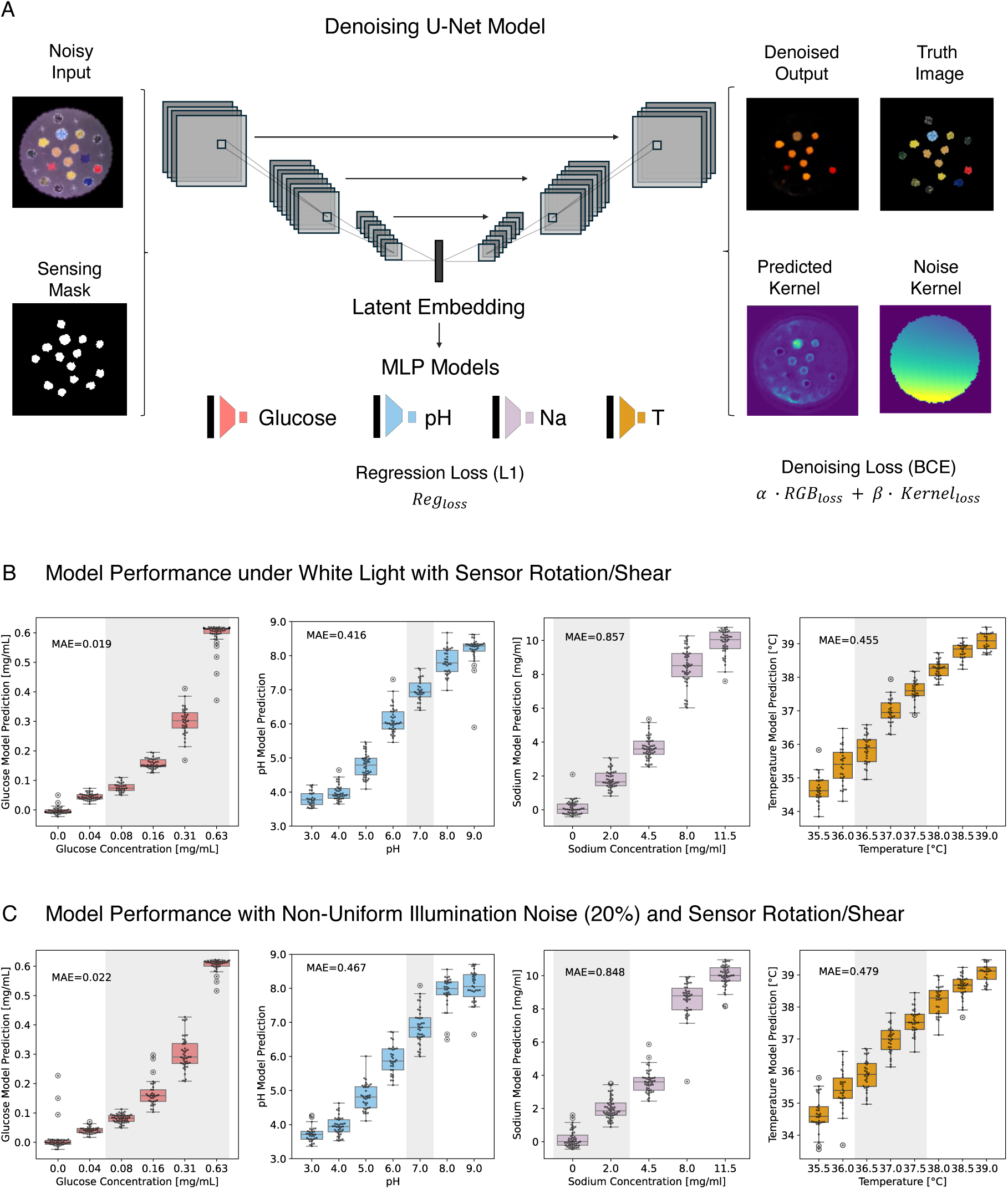
Deep Learning for Colorimetric Image-based Parameter Measurement. **A.** Developed U-Net architecture for image denoising and extraction of latent image features. Downstream parameter prediction from latent embeddings. The denoising multitask loss is formulated as image reconstruction loss (*RGBloss*) and illumination noise kernel (*Kernelloss*). **B.** Performance of DL-models for parameter prediction in white light and (128×128) resolution. **C.** Performance of DL-models for parameter prediction with non-uniform illumination noise (128×128). (Shaded areas correspond to clinically relevant thresholds for interstitial fluid).

Models were trained and tested with an augmented dataset built from the experimental color distributions of each colorimetric dye and a set of real pictures of the sensor skeleton with applied rotation and shear transformations (*Materials and Methods, c*; ***Sup. Fig. 3-5***), the color-checker references and background areas also provide cues for the illumination correction task. Additionally, two-step baseline methods that first apply a global color correction to the sensor image, based on the color checker references, and a subsequent color regression were studied [35]. As they only estimated a global color homography and can therefore not correct for spatial non-uniform illuminations [39]-[40], the two-step baseline methods showed decreased performance [35].

Performance was evaluated with sensor shear/rotation in two illumination conditions (***Fig. 3b-c***): *(i)* with a colorimetric response in standard white-light illumination, *(ii)* with perturbed illumination, using non-uniform spatial noise distributions (*Materials and Methods, c*). Prediction quality was measured as the Mean Absolute Error (MAE) and the Spearman’s rank correlation coefficient (ρ) between the image labels and predicted values.

Parameter estimation in the sensing ranges of each dye was achieved with high precision by the U-Net MLP under neutral white light for the target variables, with sensor shear/rotation and image resolution of (128×128) pixels: Temperature (MAE=0.455°C, ρ=0.968 p-value=3.23e-133); pH (MAE=0.416, ρ=0.967 p-value=2.56e-143); Sodium (MAE=0.857mg/mL, ρ=0.965 p-val=6.49e-140); and Glucose (MAE=0.0187mg/mL, ρ=0.982 p-val=1.83e-173) (***Table 1***, ***Fig 3b***).

**Table 1.** Colorimetric Reading Model Performance with Sensor Image Shear/Rotation and Illumination Noise.

| MAE<br>± SD | Colorimetric<br>Deep Learning Model | Temperature<br>[°C] | pH | Sodium<br>[mg/mL] | Glucose<br>[mg/mL] |
| --- | --- | --- | --- | --- | --- |
| White<br>Illumination | CNN + MLP<br>Regression | 0.582<br>± 0.348 | 0.343<br>± 0.367 | 2.452<br>± 2.402 | 0.050<br>± 0.041 |
|  | U-Net coupled with<br>MLP Regression | 0.455<br>±0.379 | 0.416<br>± 0.356 | 0.857<br>± 0.701 | 0.019<br>± 0.026 |
| Non-Uniform<br>Illumination<br>Noise (10%) | U-Net coupled with<br>MLP Regression | 0.519<br>±0.384 | 0.426<br>± 0.338 | 0.847<br>± 0.748 | 0.020<br>± 0.024 |
| Non-Uniform<br>Illumination<br>Noise (20%) | U-Net coupled with<br>MLP Regression | 0.479<br>±0.397 | 0.467<br>± 0.376 | 0.848<br>± 0.672 | 0.022<br>± 0.029 |

In addition, the model’s ability to detect clinically relevant pathological thresholds in neonatal interstitial fluid was assessed. The model demonstrated high balanced accuracy (b. acc.) and weighted (w.) F1-scores in identifying critical thresholds for cystic fibrosis (b. acc. 91.40%, w. F1-score 93.19%) for sodium above 2.0 mg/mL [41]; hypernatremia (b. acc. 92.52%, w. F1-score 90.95%) for sodium above 3.5 mg/mL [42]; hypoglycemia (b. acc. 98.08%, w. F1-score 99.16%) for glucose below 0.5 mg/mL [6], [10]; pH imbalances, with identification of pH above 7.5 (b. acc. 93.36%, w. F1-score 95.77%,) and pH below 7 (b. acc. 92.99%, w. F1-score 94.10%), for a healthy pH range of 7.30-7.45 [43]]; and thermal dysregulation with hyperthermia (b. acc. 96.58%, w. F1-score 97.15%) and hypothermia (b. acc. 98.11%, w. F1-score 98.17%) when outside the 36.5 – 37.5°C range [44] (See confusion matrices ***Sup. Fig. 1***, and pathological thresholds in ***Sup. Table S1***).

Colorimetric prediction by the U-Net MLP model preserved high read-out performance when introducing challenging illumination conditions (***Table 1)***. Perturbations were introduced by random color illuminations and non-uniform spatial kernels at two noise intensities, i.e., 10% and 20%. At a noise level of 20%, the model’s predictions for Temperature (MAE=0.479°C, ρ=0.9562 p-val=1.31e-129); pH (MAE=0.467, ρ=0.9564 p-val=2.93e-129); Sodium (MAE=0.848mg/mL, ρ=0.966 p-val=3.10e-141) and Glucose (MAE=0.0224mg/mL, ρ=0.964 p-val= 7.18e-139) retained sufficient accuracy, demonstrating robustness to both complex illumination artifacts and image shear/rotations (***Fig 3c***).

### C. Pilot for Clinical Application

#### i. Adaptation of Silk Colorimetric Sensor Design to Clinically Relevant Conditions

*Microfluidic evaluation:* The sensor’s design was optimized for microfluidic performance using capillary effects, enabling the simultaneous contact of multiple dyes with highly comparable fractions of perspiration fluid, while minimizing contact of the sensing dye areas with the skin. Starting from the sole bottom inlet, the fluid effectively reaches the sensing areas through a capillary net that holds 35 μl (***Fig. 4a***). This renders the sensor’s design clinically applicable, as the transepidermal fluid loss rate for preterm neonates varies between 15-22 g/m^2^/ hr [13], resulting in a transepidermal sample volume of 23.8-34.8μL (i.e. 2.1 – 3.2μl per sensing area, for n=11) extrapolation of 38 minutes, for a Tegaderm patch area of 5×5cm. In the experimental setting, we successfully showed effective colorimetric dye response using small sample volumes between 1-2μL per sensing area for pH, glucose, and sodium.

**Figure 4.**
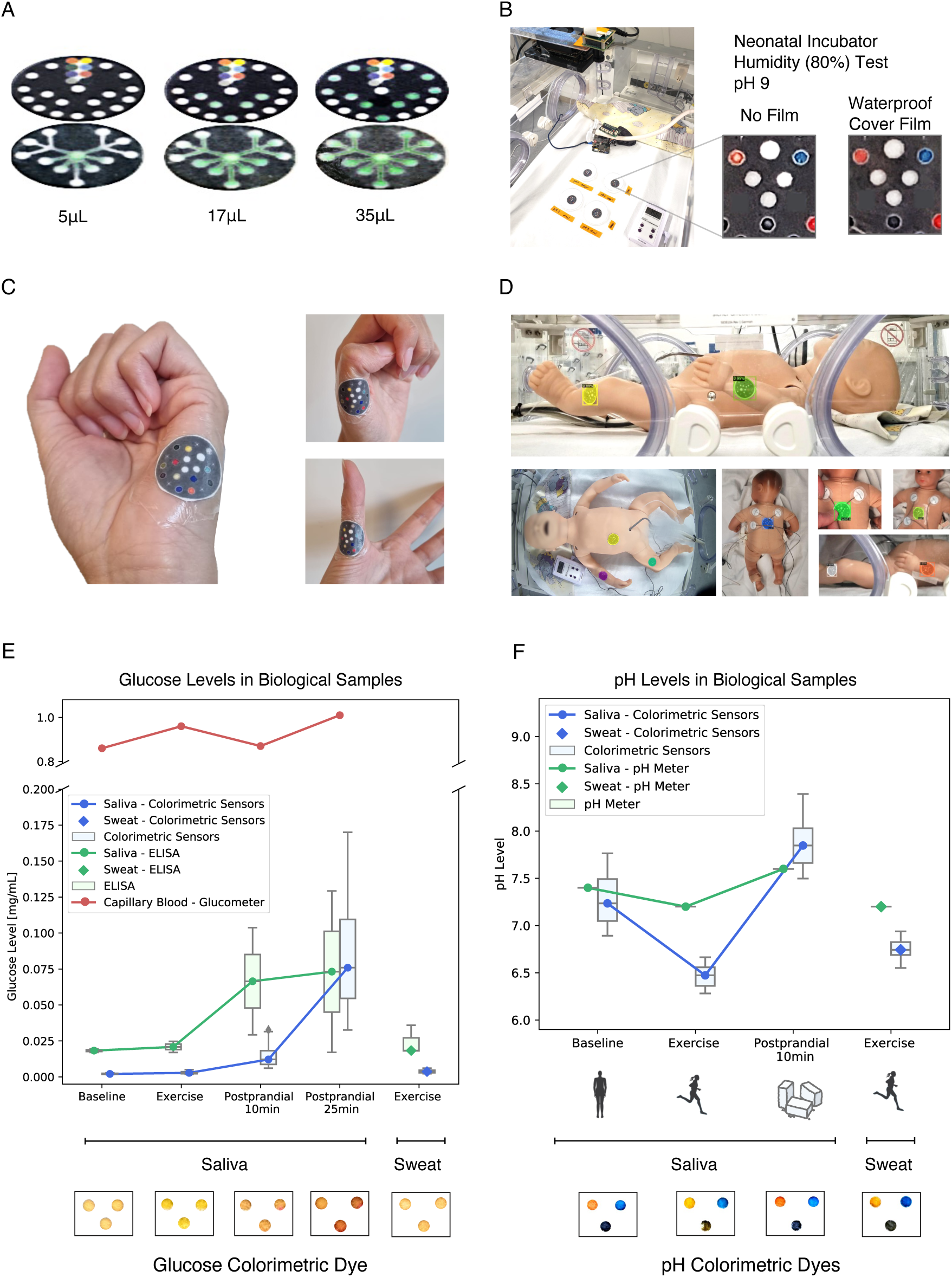
Pilot experiments for assessment of clinical application. **A.** Demonstration of passive biofluid flow using the microfluidic design total capacity of 35 μL. **B.** Incubator humidity test (80%) for validation of color saturation after 120 min exposure without (left) and with (right) a waterproof cover layer. **C.** Skin compliance and conformability exemplified by adhesion of the sensor to different hand regions using a medically approved adhesive film. **D.** Clinical conditions for video sensor detection, with examples of model predictions and image labels (bottom left). **E-F.** Pilot evaluation of glucose and pH colorimetric measurements in perspiration and saliva samples.

*Dye stability under clinical conditions:* Neonatal incubators provide a controlled environment to support critical body functions in the neonate through elevated temperature and humidity. We re-evaluated the performance of the colorimetric sensors inside the incubator (***Fig. 4b***), with 60-80% humidity and with temperature increasing from 34 to 39°C, successfully demonstrating preservation of the pH dye color saturation after 120 minutes, comparing a set of two sensors protected by medical water-proof film, against two sensors without the water-proof film. The average pH color saturation after 120 minutes was higher for the film-covered dyes (PR=67.1%, BG=81.2%, NY=25.0%), and lower without the impermeable film (PR=46.3%, BG=64.8%, NY=15.4%).

*Skin Conformability:* The use of flexible materials for the sensor (i.e., filter silk paper in combination with 3M™ Tegaderm™) optimized skin contact of the sensor, enabling conformance to body curvature, while avoiding skin traction (***Fig. 4c***).

#### ii. Automated Sensor Detection in the Neonatal Incubator

Detection and segmentation of the colorimetric sensor in a pilot video dataset obtained in the neonatal incubator showed high performance (0.986 AP @ IoU=0.50) using a *Mask R-CNN* model with a feature pyramid network (FPN) [38], [45] (*Materials and Methods, c*). The dataset reflected prevalent conditions of neonatal intensive care including patient movement, object obstruction by hands of healthcare personnel, electrodes or bedclothes, and varying high or low light conditions including shadows and incubator glass reflections (***Fig. 4d***). Performance comparisons (***Table 2***) allowed us to identify most challenging scenarios for segmentation such as obstruction of the sensor area by silver colored electrode cables.

**Table 2.** Per Category Instance Segmentation Results on the Incubator Dataset.

| Category of Video Recording | AP<sub>IOU=0.50:0.95</sub> | AP<sub>IOU=0.50</sub> | AP<sub>IOU=0.75</sub> |
| --- | --- | --- | --- |
| Patient Movement Simulation | 0.823 | 0.983 | 0.880 |
| Obstructions<br>(Hands, Bedclothes, Electrodes). | 0.695 | 0.925 | 0.730 |
| Low Illumination | 0.820 | 1.000 | 1.000 |
| Reflections in the Incubator Glass | 0.770 | 0.995 | 0.995 |
| Unseen Camera View | 0.890 | 1.000 | 1.000 |

*Best (1 Class) Mask R-CNN Model. AP=average precision. IOU=Intersection over Union.*

#### iii. Pilot Evaluation in Human Biofluid Samples

Evaluation of the pH and glucose colorimetric sensor’s performance was assessed with non-invasively collectable biological fluids, using samples of adult perspiration fluid and saliva. Samples were obtained under four conditions: fasting, post-exercise, 10 minutes, and 25 minutes post-prandial. Validation of the sensor’s measurements was obtained in parallel, using standard measurement devices, i.e., a laboratory pH meter and an ELISA glucose kit. Additionally, the capillary blood glucose levels were obtained as biological reference values for every condition (*Materials and Methods, d*).

The colorimetric sensor showed glucose concentrations below 0.003mg/ml in saliva samples during fasting and exercise, followed by an increase in glucose levels at 10 min and 25 min postprandial, estimating median glucose concentrations of 0.012 mg/mL (0.008 – 0.018 mg/mL, interquartile range (IQR)) and 0.076 mg/mL (0.055 – 0.110, IQR), respectively. The increase in glucose levels was also reflected by ELISA measurements in a sample aliquot showing median glucose concentrations of 0.018 mg/mL and 0.021 mg/mL during fasting and exercise and increasing to 0.067 mg/mL (0.048 – 0.085, IQR) and 0.073 mg/mL (0.045 – 0.101, IQR) 10 min and 25 min postprandial (***Fig. 4e)***. In samples from perspiration fluid, the colorimetric sensors indicated a concentration below 0.004 mg/ml, reflected by ELISA demonstrating low glucose concentration in perspiration fluid with 0.018 mg/mL. The blood glucose levels (capillary blood/glucose meter) confirmed an increase of glucose from 0.86 mg/mL under the fasting condition, to 0.87 mg/mL and 1.01 mg/mL for 10min and 25min post-prandial.

The colorimetric sensor showed a decrease in pH from 7.24 under fasting to 6.47 under post-exercise and an increase to 7.85 under post-prandial (10 min) in the saliva samples. The findings were confirmed in the saliva samples by pH meter with a pH of 7.40 under fasting to 7.20 post-exercise and 7.60 under post-prandial (10 min) condition. The pH levels in perspiration samples confirmed low pH values post-exercise with 6.74 (colorimetric sensor) and 7.20 (pH-meter) (***Fig. 4f)***.

## 3. Discussion

We showcase shelf-stable colorimetric sensing patches for automated non-invasive monitoring of critical body function biomarkers, via perspiration or saliva, leveraging the ability of silk fibroin to stabilize labile molecules. The sensor’s optimized design realizing multiple dyes on a miniaturized patch with a skin-friendly bottom sample inlet caters to the clinical requirements of a sensitive patient population. Automated quantification of the colorimetric response via deep learning models provided accurate and robust measurements in relevant ranges and under critical clinical conditions such as image perspective shear/rotation, uncontrolled light, and enabled sensor tracking on a moving object, key requirements for their implementation as a wearable monitoring technology in the neonatal incubator. Functionality with biological fluids was successfully demonstrated in an experimental setting with different physiological conditions.

The colorimetric dyes showed high sensitivity for parameter measurement in ranges relevant in neonatal care (***Fig 3c***), indicating the potential for clinical use to identify critical events (e.g., metabolic acidosis/alkalosis [43], glycemia [3], cystic fibrosis [41], [46], sepsis [47]) by changing levels of the respective target in interstitial fluid. Thresholds could be reliably detected, including cystic-fibrosis (93.2% w. F1-score), hypernatremia (91.0% w. F1-score), hypoglycemia (99.2% w. F1-score), pH imbalance (below and above healthy range, with 94.1% and 95.8% w. F1-score), hyperthermia (w. F1-score 97.2%) and hypothermia (w. F1-score 98.2%), thereby showing capability to support real-time clinical decision-making in critical conditions.

The microfluidic design for passive perspiration sample collection enables multiarray sensing holding a total volume of 35μL (for 11 sensing areas), realizing measurements in samples as small as 3μL per sensing area. The sample volume is comparable to the requirements of clinical iontophoresis in infants (>18μL in 30 minutes) [9], while matching the natural rate of transepidermal fluid loss of preterm neonates in 15-30 minutes [13]. The sensor thereby reaches a milestone in catering to the demand of smaller sample volumes than alternative devices for passive adult perspiration collection (50-100μL) [21], [29], [30].

Furthermore, the lightweight (0.05g) colorimetric paper sensor patch resulted in a curvilinear, highly conformal water-resistant interface needed for contacting the fragile newborn skin, facilitating the rapid attachment of the adhesive patch and amplifying substrate stability in the humid conditions of the neonatal incubator.

As a critical prerequisite for device application in real-life, clinical settings, the deep learning models showed high performance for color-based parameter reading in varied conditions, with accuracy comparable to previously published studies, including sensor shear (acc. 89%)[25] and fixed illumination setups (acc. 99%) [24]. Moreover, accurate segmentation of the sensors in videos recorded inside the neonatal incubator was achieved, including high prevalence conditions such as object movement, varied illumination, and the appearance of occlusion elements, allowing identification of challenging edge cases (e.g., silver-colored electrodes) for future model fine-tuning. Conditions of the daily clinical workflow can critically limit the technology performance, as observed in studies using computer vision models in the neonatal intensive care unit (NICU) [48], and therefore require extensive evaluation for transition into the clinic.

The proposed deep learning models for colorimetric read-out can be adapted to other sensor designs, requiring only the sensor mask and representative color distribution examples for each parameter (*see* https://github.com/SchubertLab/biofluid_colorimetric_sensors). Similarly, the proposed data augmentation pipeline, which accounts for sensor shear and non-uniform illumination, is transferable to other point-of-care imaging applications that involve uncontrolled lighting conditions.

Pilot validation measurements in biological fluids successfully reflected the pre– and post-prandial variation of glucose levels, as well as physiological changes to pH upon exercise, demonstrating the capacity of the colorimetric sensor to capture physiological conditions in a real-world scenario. The observed interrelated response of perspiration fluid and capillary blood levels underlines the relevance of non-invasive biofluid measurements for prompting timely clinical interventions. In previous studies, a close relation of changes in biomarkers from saliva, perspiration, or interstitial fluid, and systemic changes in the blood have been observed, with specific examples for glucose, pH, electrolytes [9], [43], [49], [50], and successful applications in the newborn clinical setting [10].

Next steps to implement the sensor into clinical care would require parallel sampling of transepidermal fluid and capillary blood specimens at various gestational age levels to account for potential effects of newborn skin maturation, as well as the performance of measurements in different (patho)physiologic conditions to expand biomarker interpretability. Colorimetric dye optimization can include targeting additional parameters such as CO2 or oxygen saturation, and the advancement of microfluidic designs for longitudinal measurements [31].

Regarding model performance, expansion of the image dataset to encompass diverse NICU environments (e.g., different neonatal incubators, additional camera angles, and illumination conditions) across multiple medical centers, along with the incorporation of patient models that represent a wide range of skin tones and body types, is required for transition into the clinical settings. Optimization of the proposed deep neural architectures for image-based parameter prediction would be particularly needed for conditions of limited hardware resources.

Automated colorimetric sensing using the proposed sensor and AI models, in combination with the future steps outlined above, has significant potential to advance the technology into clinical care by the use of a clinical trial. The noninvasive detection of metabolic or respiratory imbalances in the most vulnerable patients can enable faster treatment modifications when needed, and/or prompt the timely collection of confirmatory and more informative blood samples.

## 4. Conclusions

The advancement of non-invasive monitoring via shelf-stable, paper-based colorimetric disposable sensors optimized for clinical needs in high-end patients in combination with automated AI-based read-out holds significant potential for transforming neonatal clinical care. By providing noninvasive monitoring for critical metabolic and respiratory biomarkers through a wearable device in perspiration or saliva, their clinical application will reduce the need for frequent invasive measurements, decrease the incidence of undetected health imbalances, and thereby improve clinical decision-making. The novel medical technology offers a low-cost, easily applicable, miniaturized solution, delivering critical information at the point of care, thereby aligning with the emerging landscape of digital health technologies while expanding access to high-quality care in the neonate. These sensors hold significant promise for future applications in low-resource or underserved settings.

## 5. Materials and Methods

### A. Sensor Fabrication and Design

*Materials:* Sodium carbonate, lithium bromide, Glucose Oxidase from Aspergillus niger Type VII (Gox), Peroxidase from horseradish Type I (HRP), sodium 3,5-dichloro-2-hydroxybenzenesulfonate, 4-aminoantipyrine, chitosan, acetic acid, bromocresol green sodium salt, nitrazine yellow, phenol red sodium salt, chromoionophore I, sodium ionophore X, potassium tetrakis(4-chlorophenyl)borate, poly(vinyl chloride), bis(2-ethylhexyl) sebacate, tetrahydrofuran, and Whatman™ Grade 1 filter paper were purchased from Sigma-Aldrich (USA). Acid Yellow 34 was purchased from Chem Cruz (USA). Thermochromic pigments were purchased from Atlanta Chemical (USA). Transparent double-sided adhesive films were provided by FLEXcon (USA). Tegaderm™ film dressings (size: 4.4 x 4.4 cm) were purchased from 3M (USA). Silk cocoons of *Bombyx mori* silkworm were purchased from Tajima Shoji (Japan). Wax ink blocks were purchased from Xerox (USA). Deionized (DI) water with resistivity of 18.2 MΩ cm was obtained with a Milli-Q reagent-grade water system and used to prepare aqueous solutions.

*Silk fibroin solution preparation:* Silk fibroin was extracted following a previously reported protocol. Briefly, finely chopped *Bombyx mori* silk cocoons were boiled in a solution of 0.02 M sodium carbonate to remove the sericin layer for 120 minutes. The fibers were washed three times for 20 minutes in DI water, dried overnight, and dissolved in a solution of lithium bromide 9.3 M at 60 °C for 4 hours. The obtained solution was dialyzed against deionized water for 2 days, changing the deionized water 6 times at regular intervals. The final solution was centrifuged twice at a speed of 9000 rpm, at 4 °C, for 20 minutes and then filtered, yielding 7-8 wt% silk fibroin solution.

*Chromogenic Enzymatic Glucose-sensing areas*. Silk-based chromogenic enzymatic inks were prepared using 4 wt% silk fibroin solutions containing 0.1 M PBS to maintain a stable pH when reacting with strongly acidic or basic sweat. Enzymatic reagents were added to the silk solution at final concentrations of 339 U mL⁻¹ for horseradish peroxidase (HRP) and 645 U mL⁻¹ for glucose oxidase (GOx). Chromogenic substrates were then dissolved in the silk-based enzymatic mixture to yield final concentrations of 0.75 mg mL⁻¹ of 4-aminoantipyrine, 0.3 mg mL⁻¹ of acid yellow 34, and 1.7 mg mL⁻¹ of sodium 3,5-dichloro-2-hydroxybenzenesulfonate. The circular sensing areas were first functionalized with 0.4 µL of a chitosan solution consisting of 0.5 wt% chitosan dissolved in 2 wt% acetic acid, which was allowed to dry at room temperature. Subsequently, 0.4 µL of the chromogenic enzymatic ink was drop-cast onto the chitosan layer and allowed to dry at room temperature. This deposition process of the silk-based ink was repeated four times to obtain a total of four layers of ink.

*Chromogenic pH-Sensing Areas*. Silk-based chromogenic pH-sensitive inks were made of 4 wt% silk fibroin solutions containing a pH indicator (i.e., phenol red sodium salt, nitrazine yellow, bromocresol green sodium salt) with a final concentration of 2.5 mg mL−1. 0.3 ul of the ink was drop-cast of the circular areas. The ink was let dry at room temperature and the process was repeated a second time to obtain 2 layers of ink.

*Chromogenic Temperature-Sensing Areas*. Temperature-sensitive inks were made of 65 wt% DI water, 32.5 wt% Mod Podge and 2.5% thermochromic pigment (Atlanta Chemical, USA). 0.5 ul of the ink was drop-cast of the circular areas. The ink was let dry at room temperature and the process was repeated a second time to obtain 2 layers of ink.

*Chromogenic Sodium-Sensing Areas*. A sodium-sensing ink was prepared consisting of chromoionophore I (0.06 wt%), sodium ionophore X (0.12 wt%), potassium tetrakis (4-chlorophenyl)borate (0.11 wt%), poly(vinyl chloride) (0.86 wt%), bis(2-ethylhexyl) sebacate (3.43 wt%), and tetrahydrofuran (95.41 wt%). The mixture was prepared by sequential addition of the components and stirred overnight before use. The circular sensing areas were first functionalized with 0.7 µL of a 4 wt% silk solution, which was allowed to dry at room temperature, followed by 1 µL of 4 wt% silk solution to form a two-layer base. After drying, 0.1 µL of the sensing ink solution was drop-cast seven times onto each circular area on top of the silk base layer. A second 4 wt% silk fibroin solution was then applied as a capping layer by drop-casting 1 µL followed by 1.5 µL on top of the membrane layer.

Before assembling the patch, a buffer was added to the circular areas in correspondence with the sodium-sensing regions on the first paper layer of the patch that contains the flow channels. The buffer was prepared using 1 M HEPES solution adjusted to pH 6.89 (target range 6.85– 6.95) by titration with 10 M KOH. 0.5 µL of the buffer solution was drop-cast five times on the circular areas to complete the intermediate buffering layer.

*Sensor Fabrication:* Whatman™ Grade 1 filter paper was used as a substrate to fabricate the paper-based sensors. A vector graphic software (Adobe Illustrator) was employed to design the distribution pattern of the sensing areas and the fluidic flow channels. The wax pattern was printed directly onto the filter paper using a wax printer (Xerox ColorQube 8570). Afterwards, the wax-printed paper was heated at 150 °C for 30 s to allow the wax to melt and penetrate into the filter paper to create a hydrophobic channel pattern.

*Sensor Design.* A base layer of Tegaderm wound dressing film was laser cut to obtain a circular opening (3 mm) to be used as fluid inlet channel. The Tegaderm film is used as a base layer on which the sensor can easily be attached and detached to minimize skin discomfort. The sensing patch consists of two wax-printed paper layers: the first layer consists of a ramification of flow channels that transport the fluid from the inlet opening to the sensing areas, and the second layer consists of the sensing areas and the color references. The two layers are attached using adhesive films.

### B. *In vitro* Validation

Experimental validation of the dye colorimetric response was performed on (n=34) silk-fibroin sensors under white light conditions.

*Temperature Calibration:* We designed a standardized experimental setup to capture video recordings of the colorimetric response to temperature (***Sup. Fig. 6b***; Video: 30fps, ultra-lowlight 4K Camera with 8 MP (*e-CAM80_CUNX*), Image Sensor: *Sony 4K STARVIS™ IMX415 CMOS,* Data collection: *NVIDIA Jetson Xavier NX Board*). Sensors (n=3) were positioned between impermeable Tegaderm films and secured to the inner surface of a glass container, oriented towards the camera for optimal visibility. The container was filled with water, and a closed-loop control heater was employed to maintain precise temperature regulation. The colorimetric responses were recorded simultaneously as the water temperature was increased from 31°C to 42°C every 0.2°C in 1-minute intervals, as well as upon return to room temperature (42 to 30°C, n=3) to measure hysteresis.

*pH Calibration:* Standardized video recordings captured the color response of two sensors (n=2) to solutions with varying pH (***Sup. Fig. 6c***). While a pH-meter (*WTW inoLab pH 720*) provided real-time pH measurements, acidic (*HCl 37%*) or basic (*NaOH 1M*) solutions were incrementally added to distilled water in a beaker under constant homogenization through a magnetic stirrer. The pH was increased in increments of 1 pH unit every 5 minutes in the range from pH=3 to pH=9, before reversing from 9 to 3 in identical steps. The sensors were in direct contact with the liquid solution from the top of the beaker, positioned on a plastic mesh device. The cyclic pH experiments, i.e., capturing of bidirectional colorimetric changes showed a color saturation loss of 28% as a consequence of the required water submersion for the experimental setup that had to operate without the waterproof film cover. In addition to the cyclic experiment, images of seven different sensors (n=7) were captured through a smartphone camera (*12 MP/64MP, Galaxy S21*) while directly applying 3μL of solutions with a pH of 3, 4, 5, 6, 7, 8, and 9 to the bottom inlet of the pH sensing area.

*Sodium Calibration:* Images of (n=11) sensors were captured (*12 MP/64MP, Galaxy S21*) while applying 3μL standard solutions containing 50, 100, 130, 140, 150, 180, 200, 300, 500, 700, and 1000 mmol/L sodium chloride (NaCl) to the bottom inlet of the sodium sensing area. *Glucose Calibration:* The colorimetric response for glucose was evaluated from pictures (*12 MP/64MP, Galaxy S21*) of (n=8) sensors exposed to 2μL of solutions with decreasing concentrations of glucose, following a serial dilution protocol. The glucose concentrations were 10, 5, 2.5, 1.25, 0.625, 0.3125, 0.1563, 0.078 mg/mL, and distilled water.

*Methods for color calibration:* Images for *in vitro* validation were white balanced by using the color checker references and applying color level adjustments (matching the white and 50% grayscale) with the software *GIMP 2.10*. Our computational imaging pipeline was applied for three tasks: (1) locate the sensor, (2) register, segment, and label the corresponding colorimetric areas, and (3) extract the pixel values of the sensing area (***Sup. Fig. 6a***, see the code here: *github.com/SchubertLab/biofluid_colorimetric_sensors*). For quantification of the colorimetric response, sensitivity was calculated from the slope of the linear effective sensing range from the calibration curves (***Fig. 2***).

### C. Deep Learning Imaging Methods

*Pipeline for Experimental Data Augmentation:* Images of the sensors under white light (scene illuminated with LED 4500K) were used to capture the pixel color distributions for each of the variables (***Sup. Fig. 3***). Pictures of multiple sensor skeletons (*n=20*) were obtained from the experimental *in-vitro* validations, and sensing areas were masked. Random pixels from the experimental color distributions were sampled into the corresponding sensing areas of a sensor skeleton, generating augmented sensor images that contain different combinations of all the measured parameters (pH, temperature, glucose, sodium) at randomly sampled levels.

*Illumination Perturbations:* The data generation pipeline randomly sampled illumination noises with different spatial distribution kernels *f*(*x*, *y*), with random linear, polynomial, and radial distributions [35], adding them to every color channel (*R_ch_*, *G_ch_*, *B_ch_*) with a different intensity value (α, β, γ), bound from 0 to ±10% or ±20% of the overall intensity (0-255), (***eq. 1***). After the noise kernel was added to each RGB channel, a min-max image normalization was applied. Image rotation (±20°), shear (±0.1%), and random cropping (±0.1%), as available in torchvision transforms, were added to simulate real test conditions and generated a final noisy image *I_noise_*.

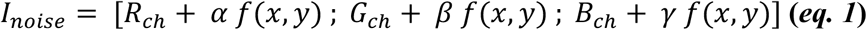

*Colorimetric Parameter Reading – Baseline Model:* Estimation of parameters from the sensor images under reference white light (scene illuminated with LED 4500K), including image zoom/shear/rotation, was first performed with a deep learning model for every variable, consisting of a convolutional neural network (CNN) coupled with an multilayer perceptron (MLP) model, the model input consisted of the RGB sensor image and a mask of the target sensing areas, with an input size of (128,128,4). The ground truth label was the corresponding variable value min-max normalized in the measured range from [0–1].

The CNN module consisted of two convolutional layers (32 filters and 64 filters, respectively, both with a kernel of 5), followed by max pooling and ReLU operations. After applying a flattening operation, the MLP module consisted of 3 fully connected layers (with 53824, 2600, and 1200 neurons), followed by a ReLU operation. The last neuron integrated a sigmoid activation function. The final model configuration was the result of a hyperparameter optimization using Optuna [51], aiming to minimize the mean square error of the predictions in the validation set. We also tested prediction performance with a RESNET18 module (testing both with pretrained weights or untrained) as a feature extractor, which showed lower performance. Different activation functions and learning rates were also evaluated (see final values of the hyperparameters in ***Sup. Table III***).

A total of 3200 images were used for training/validation of each CNN+MLP model for parameter regression, with a total of 20 epochs (128 training images and 32 validation images per epoch). Evaluation was performed on 240 images for every parameter.

*Colorimetric Parameter Reading and Color Correction – U-Net Model:* Estimation of the parameters from sensor images under varying non-uniform illumination noise, was conducted by developing denoising autoencoders based on the U-Net model [38] coupled with MLPs to perform regression for each variable from the latent features, thus integrating both the color correction and inference tasks into a single framework (*code available in: github.com/SchubertLab/biofluid_colorimetric_sensors*). The input consisted of the noisy RGB image and a fourth channel with a mask of the relevant sensing areas, with input size (128, 128, 4); the models were then trained to disentangle the illumination noise, by reconstructing the original RGB image colors (only with the sensing areas), and an estimation of the spatial distribution of the illumination noise on the image (***Fig. 3a***). The loss of the denoising autoencoder (*D*_*loss*_) was therefore defined as the sum of the losses for each RGB channel and the illumination kernel loss (*K*_*loss*_) (eq. 2). The best U-Net denoising loss combined a sigmoid function and a Binary Cross Entropy loss, and was implemented as a pytorch function *BCEWithLogitsLoss (pytorch 2.2.2)*.

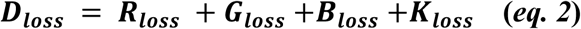

Simultaneously, the U-Net latent features were passed to the MLP networks as input, the MLP models were trained with a L1 loss between the parameter predictions and the corresponding label for the sampled variable. Labels were min-max normalized in the sensing range from [0–1] for every variable in their measurement range.

The denoising U-Net architecture comprised four encoding blocks, each containing two convolutional layers, followed by batch normalization and ReLU activation functions. The first encoding block utilized 32 filters, with the number of filters increasing progressively in subsequent blocks by factors of 2, 4, and 8, respectively. This hierarchical encoding resulted in a latent representation with a dimensionality of 32768 neurons. The decoding path mirrored the encoder’s structure, consisting of four up-convolutional layers. Each decoding stage incorporated skip connections from the corresponding encoding layer to facilitate spatial information recovery and enhance feature propagation. The MLP latent regression module consisted of 4 fully connected layers (the size corresponding to the latent dimensions divided by 4, 8, and 16, respectively). The last neuron had a sigmoid activation function. See model and training scripts in (http://github.com/SchubertLab/biofluid_colorimetric_sensors). Evaluation with other loss types and learning rates was performed to select the optimal model configuration; the chosen configuration had the lowest MSE in the validation set, see final hyperparameter values in ***Sup. Table IV***.

A total of 9600 sensor generated images were used in training and 2560 for validation of the U-Net MLP model (40 epochs, 240 train images and 64 validation images per epoch); evaluation is performed on 240 sensor images for every parameter.

*Sensor Segmentation in the Incubator:* An instance segmentation model using a *Mask R-CNN* model with a feature pyramid network (FPN) [38], [45], was trained for real-time detection of the newborn wearable sensors in the neonatal incubator conditions. Models were trained and tested on challenging conditions such as movement of the infant, temporary occlusions (Sheets, hands, electrocardiography electrodes), and varying illumination (see ***Sup. Table. II***). During training, input images were resized to 800×1333 pixels and augmented via contrast and saturation jittering, as well as random rotations. The Region Proposal Network (RPN) used anchors at five scales and three aspect ratios. The model was trained for 7000 iterations using Adam (learning rate=4.5×10^-3^, decayed by 0.1 at iterations 5000 and 6000; weight decay=1×10^-4^; momentum=0.9). The model architecture was optimized through hyperparameter tuning using Optuna [51] (see ***Sup. Table V***).

At inference, the top 1000 proposals were retained from the FPN stage, bounding-box regression was applied, followed by non-maximum suppression. The mask branch was then executed on the 100 highest-scoring detections. For each region of interest (ROI), only the mask corresponding to the predicted class was selected and resized to the ROI’s dimensions. The predicted masks were binarized at a 0.5 threshold to produce the final segmentations.

### D. Pilot Clinical Experiments

*Sensor Detection in the Neonatal Incubator:* Videos of the sensors in a neonatal incubator (*Giraffe OmniBed, GE Healthcare*) were recorded at the Perinatal Center of the LMU Hospital in Munich. The sensors were attached to different body parts (chest, back, arm, and leg) of two baby mannequins (***Sup. Fig. 7***). One of the mannequins (*SimNewB, LaerdalMedical, Puchheim, Germany*) was equipped with a mechanical ventilation system to simulate chest movements at different breathing rates (30, 60, 90 breaths per minute) and automated limb motion (arms and legs). The second mannequin was manually placed in different body positions. A setup with the embedded system and camera (*NVIDIA Jetson Xavier NX Board, e-CAM80_CUNX 8MP*) was fixed on top of the incubator, while a smartphone camera was used on the side of the incubator (*12MP/64MP, Samsung Galaxy S21*). The collected dataset consisted of 31 videos under different illumination conditions (i.e., white LED room light, ambient natural light, low light with an incubator blanket cover) as well as under simulation of conditions as typically encountered in the clinic (i.e., cardiac electrodes attached, hands of clinical staff, partial occlusion of the sensor with bed sheets, reflections from a plastic cover; ***Sup. Table. II***). Video recording was performed for 30-180 seconds. Videos were subsampled to extract frames for manual labeling using the annotation tool for semantic segmentation *LabelMe* [52]. A total of 655 images (15 or 25 frames per video) were annotated by six trained researchers, highlighting the location of the sensor(s) in the image to subsequently train and test image segmentation models (Train/test split per scenario ***Sup. Table. II***).

*Dye stability testing in the neonatal incubator:* The saturation of the colorimetric dyes was tested inside a neonatal incubator (*Giraffe OmniBed, GE Healthcare*) while applying high humidity of 80%, and temperatures of 37-39°C. Water solutions with a pH=9 were added to cotton pads using a pipette. Two cotton pads were coated with 500 μL, and two pads with 1000μL of the solution. Sensors (n=4) were placed on top of each pad; two of them were covered with a waterproof top film (*Tegaderm, 3M*), while the remaining two were left without the impermeable top-film. The colorimetric sensors were recorded with a video camera for 120 min, placed on top of the incubator (*NVIDIA Jetson Xavier NX Board, e-CAM80_CUNX 8MP*), the humidity and temperature inside the incubator was confirmed using a coral board environmental board and coral board (*Google Coral Environmental Sensor Board V1.1, Coral Development Board*) (***Sup. Fig. 6d***). Color values were digitized with our Python video processing pipeline.

*Evaluation with biological fluid samples:* Biological fluid samples (i.e., perspiration fluid and saliva) were collected from an adult female individual in four conditions: fasting, post-exercise (20 minutes of cardio exercise), 10, and 25 minutes post-prandial. Human biological fluid samples were obtained through the CPC-M bioArchive at the Comprehensive Pneumology Center (CPC Munich, Germany). The study was approved by the local ethics committee of the Ludwig-Maximilians University of Munich, Germany (Ethic vote #19-629). Written informed consent was obtained from the study participant.

For every condition, one saliva sample (*1ml*) was collected for analysis with the colorimetric sensors, and a validation measurement was performed with an enzyme-linked immunosorbent assay (ELISA) for glucose measurement, paralleled by a glucose-meter measurement in capillary blood (*ACCU-check Performa, CE 0088, Roche*). Post-exercise, an immediate perspiration sample (*0.5ml*) was collected from the forehead and back of the participant for pH measurement. The pH of the saliva and perspiration samples was validated using a laboratory pH meter (*WTW inoLab pH 720*).

Colorimetric sensor measurements for glucose and pH were achieved by bringing the perspiration or saliva samples, respectively, to the bottom inlet of the colorimetric sensors using a volume of 3μL per sensing area. Serial images of the sensors were captured in daylight conditions with a smartphone camera (*12MP/64MP, Samsung Galaxy S21*), next to a color checker card for digital white balancing.

The parallel measurement of the glucose levels in an aliquot of the biological fluid sample was performed using an ELISA Glucose Colorimetric Detection Kit (*reference EIAGLUC, standard glucose range 0.5–32 mg/dL, <u>Thermo Fisher</u>, Invitrogen™*) following manufacturer’s instructions. Absorbance was measured with a microplate laboratory spectrometer (*SpectraMax i3x, Molecular Devices*). Glucose concentrations of the biosamples with three technical replicates were calculated by fitting a 4-degree polynomial to the standard logarithmic absorbance of the manufacturer’s references.

## Acknowledgment and Funding

The present study was supported by the Young Investigator Grant NWG VH-NG-829 (Helmholtz Association (Federal Ministry of Education and Research in Germany (BMBF)) and Helmholtz Munich, Germany) and the German Centre for Lung Research (DZL) (BMBF)). The present study is part of the transregional collaborative research center PILOT (TRR 359 Perinatal Development of Immune Cell Topology, Project number 491676693) funded through the Deutsche Forschungsgemeinschaft (DFG, German Research Foundation). A.C. is supported by the Helmholtz Association under the research school Munich School for Data Science. Computational resources for deep learning model training were supported by the BMBF-funded de.NBI Cloud within the German Network for Bioinformatics Infrastructure (de.NBI). Additional financial support was provided by the Stiftung AtemWeg (LSS AIRR).

## Data Availability Statement

The data and code repository that support the findings of this study are openly available at https://github.com/SchubertLab/biofluid_colorimetric_sensors and in the supplementary material of this article.

