## Supplementary material for "Wireless Colorimetric Multi-Biomarker Sensing to Enable Critical Neonatal Monitoring": Sup.

**Sup. Table I -** Reference values for targeted physiological variables for neonatal biological fluids (Perspiration / Interstitial Fluid or Saliva).

| **Physiological Variable** | **Perspiration or Interstitial Fluid** | **Saliva** |
| --- | --- | --- |
| pH | Interstitial Fluid - Healthy Range:  7.35–7.45 pH [43] | Healthy Range^*^:  6.2 - 7.6 pH [49] |
| Glucose | Interstitial Fluid - Healthy Range:  2.6 - 15 mmol/L [4]  2.2 - 8 mmol/L [6] | Healthy Range^*^  0.23 - 0.38 mmol/L [49]  Diabetic:  0.55-1.77 mmol/L [49] |
| Sodium | Healthy Range:  17.67 ± 7.58 mmol/L (Mean ± SD) [41]  22 mmol/L [46]  Cystic Fibrosis:  95.62 ± 28.38 mmol/L (Mean ± SD) [41]  89 (IQR 67–109)[46]  Hypernatremia (electrolyte measurements in Serum): 0.155 mol/L (3.5 mg/mL) [42] | Healthy Range:  14.37 ± 6.50 mmol/L (Mean ± SD) [9]  Cystic Fibrosis:  21.09 ± 9.29 mmol/L [41] |

**^*^**From adult studies. SD = Standard Deviation. IQR=Interquartile Range.

***Sup.*** ***Table.*** **II -** Videos Dataset for Sensor Segmentation in the Neonatal Incubator.

| **Mannequin Type** | **Camera**  **Location** | **Recording Conditions** | **Illumination** | **Sensor(s) Location** | **Train/Test Dataset** |
| --- | --- | --- | --- | --- | --- |
| Regular Baby Mannequin  (14 videos) | Top of the Incubator | Mannequin in Different Body Orientations and Rotation | Room LED white light | Chest | Train (N=25)  Test (N=25) |
|  |  |  | Natural Daylight | Chest | Train (N=25)  Test (N=25) |
|  |  |  | Low lights and Shadows (with incubator cover) | Chest | Train (N=25)  Test (N=25) |
|  |  |  | Low lights and Camera In/Out of Focus | Chest | Train (N=25) |
|  |  | Sheets/ Hand Occlusions | Room LED white light | Chest,  Back | Train (N=25)  Test (N=25) |
|  |  | ECG Cable and Hand Occlusions | Room LED white light | Chest,  Back | Train (N=25)  Test (N=50) |
|  |  | Plastic Film Reflections and High Exposure | Room LED white light | Chest | Train (N=25)  Test (N=25) |
| Mechanical Baby  Mannequin  (16 videos) | Top of the Incubator | Mannequin breathing  (30, 60, 90 breaths per minute) | Room LED white light | Chest | Train  (N=45) |
|  |  | Mannequin breathing  (30, 60 breaths per minute) and limb movement |  | Chest, Arm,  Leg | Test  (N=30) |
|  |  | Mannequin breathing  (60, 90 breaths per minute) and limb movement | Natural Daylight | Chest, Arm,  Leg | Train  (N=30) |
|  | Side of the Incubator | Mannequin breathing  (30, 60 breaths per minute) and limb movement | Room LED white light | Chest, Arm,  Leg | Train  (N=30) |
|  |  | Mannequin breathing  (90 breaths per minute) and limb movement |  | Chest, Arm,  Leg | Test  (N=15) |
|  |  | Mannequin breathing  (90 breaths per minute) and limb movement | Natural Daylight | Chest, Arm,  Leg | Test  (N=15) |
|  |  | Mannequin breathing  (60 breaths per minute) and limb movement | Low lights (with incubator cover) | Chest, Arm,  Leg | Train  (N=15) |
|  |  | Mannequin breathing  (90 breaths per minute) and limb movement | Low lights (with incubator cover) | Chest, Arm,  Leg | Train  (N=15) |
|  | New Camera View (Top right/left corners in the incubator). | Mannequin breathing  (90 breaths per minute) and limb movement | Room LED white light  (with glass reflection). | Chest, Arm,  Leg | Train  (N=15)  Test  (N=45) |

N = number of images with all sensor areas manually annotated.

**SUPPLEMENTARY TABLE III:** Hyper-parameters search ranges and best values for the CNN+MLP color-based regression model.

| **Hyper Parameter Name** | **Range / Options** | **Best Value** | **Description** |
| --- | --- | --- | --- |
| Model Type | CNN or RESNET18 | CNN | Model for Extraction of Visual Features. If RESNET18 was chosen the options (pretrained or not pretrained) were evaluated. |
| Base LR | [1e-7, 1e-2] | 7.952e-4 | Learning rate |
| Dropout | [True, False] | False | Apply 0.05 droput after every CNN layer. |
| Last Neuron Activation | [sigmoid, softplus, ReLU] | sigmoid | Activation function of the last neuron in the CNN+MLP model. |

**SUPPLEMENTARY TABLE IV:** Hyper-parameters search ranges and best values for the U-Net denoising, coupled with a latent MLP color-based regression model.

| **Hyper Parameter Name** | **Range / Options** | **Best Value** | **Description** |
| --- | --- | --- | --- |
| Model Type | Denosing U-Net or Autoencoder | U-Net | Model for performing the denoising task and extraction of latent features. |
| Denoising U-Net Loss | [BCE, MSE] | BCE | Loss for training the Denosing U-Net |
| Denoising U-Net LR | [1e-7, 1e-1] | 0.5 | Initial learning rate with a reduction to 0.05 (10% of initial value) after 20 epochs. |
| Latent MLP LR | [1e-4, 1e-2] | 1e-2 | Learning rate for latent MLP model |
| Latent MLP Neuron Activation | [sigmoid, softplus, ReLU] | sigmoid | Activation function of the last neuron in the latent MLP model. |
| Latent MLP Loss | [L1, MSE] | L1 | Loss function for training latent MLP model. |

**SUPPLEMENTARY TABLE V:** Hyper-parameters search ranges and best values for the RCNN-based sensor detection and segmentation model.

| **Hyper Parameter Name** | **Range** | **Best Value** | **Description** |
| --- | --- | --- | --- |
| Freeze Stage | [0, 5] | 2 | Backbone Freeze Stage.  0 means all the backbone is trainable, 5 means  all backbone stages are frozen. |
| Base LR | [1e-6, 1e-1] | 4.57e-5 | Learning rate |
| Backbone Length | 50, 101 | 50 | Feature Extractor Length |
| Detection Per Image | [5, 10] | 9 | Top K scoring bounding boxes considered for final prediction |
| RoI Head Threshold Score | [0.5, 0.9] | 0.68 | Threshold at which the RoI is considered as Positive prediction |
| Batch size | 2, 4, 8, 16, 32 | 16 | Mini-batch size |

**Supplementary Figure 1.**

Confusion matrices for newborn disease classification using relevant biomarker thresholds (label 0 is healthy, and label 1 is unhealthy). **A.** Hypernatremia (for sodium above 3.5 mg/mL in perspiration fluid) **B.** Cystic fibrosis (for sodium above 2.0 mg/mL in perspiration fluid). **C.** Hypoglycemia (for glucose below 0.5 mg/mL in interstitial subcutaneous fluid). **D.** pH imbalance (below 7) **E.** pH imbalance (above 7.5) , for a healthy pH range of 7.30-7.45 in interstitial fluid. **F.** Hypothermia (temperature below 36.5°C). **G.** Hyperthermia (temperature above 37.5°C).

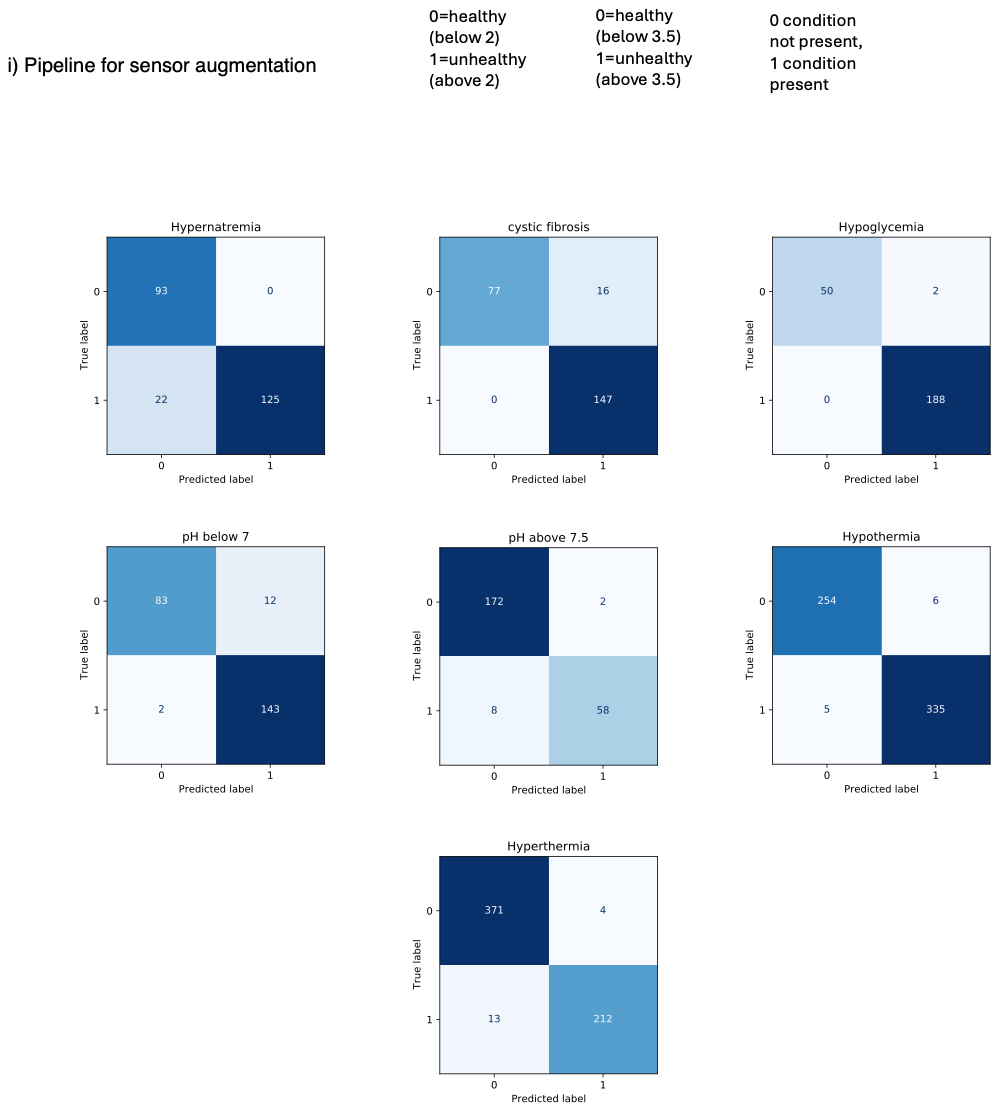

**Supplementary Figure 2.**

**A.** Temperature dye colorimetric cyclic response (down-up-down cycle).

*i. Normalized Grayscale vs Temperature*

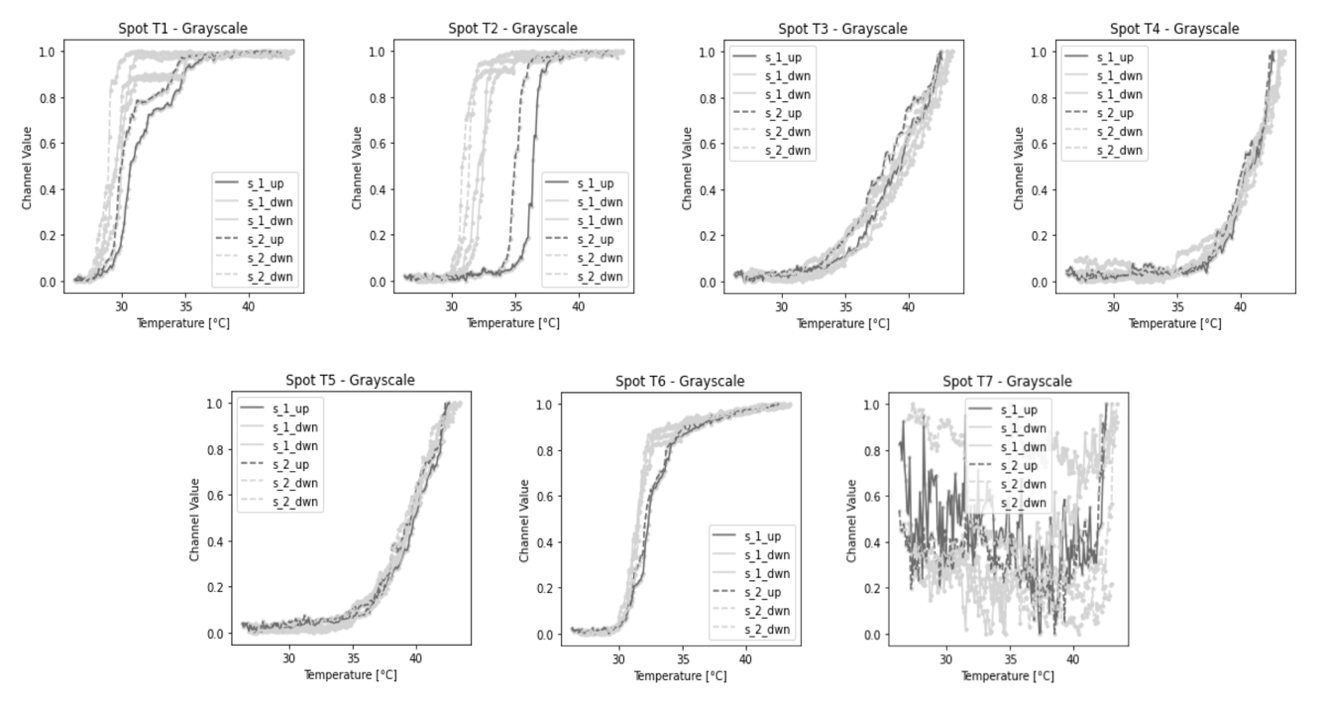

*ii. Cyclic experiment, temperature vs time. Data points display the recorded dye color.*

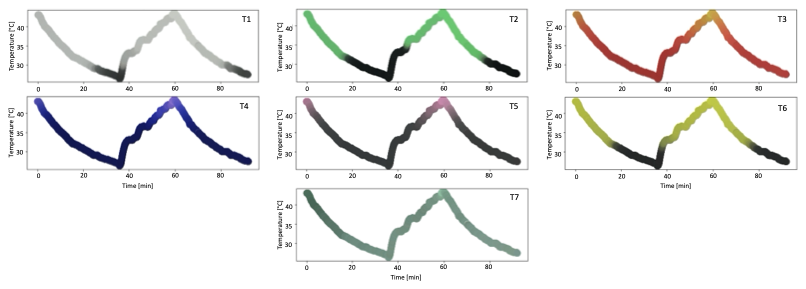

**B.** Colorimetric cyclic response for the pH dye (up-down cycle).

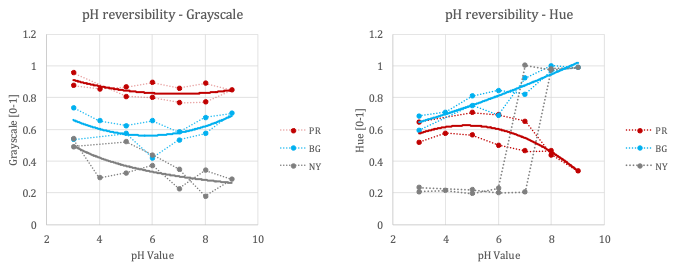

**Supplementary Figure 3**

**A.** Sensor augmentation from experimental data.

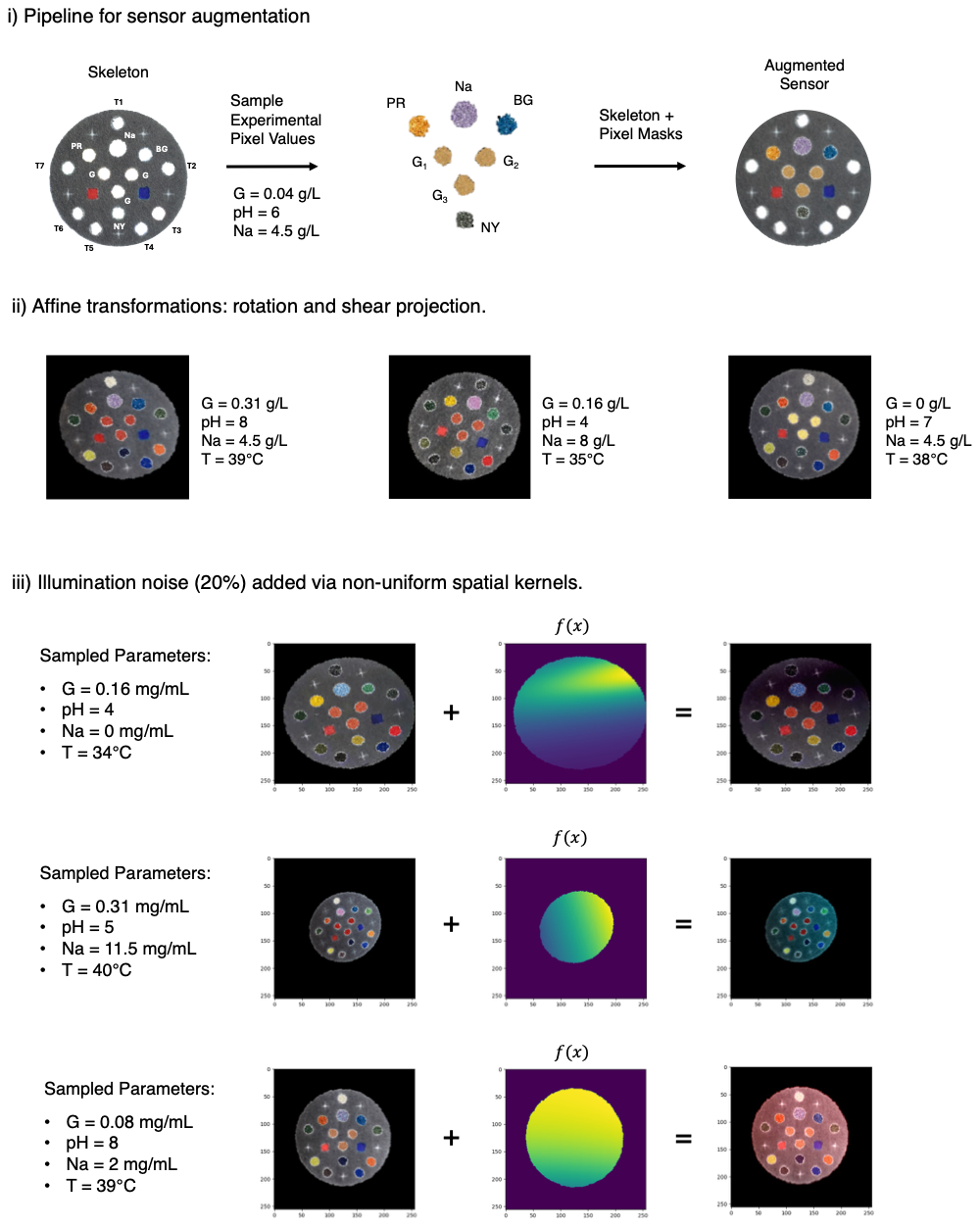

**B.** Affine transformations (rotation and shear projection).

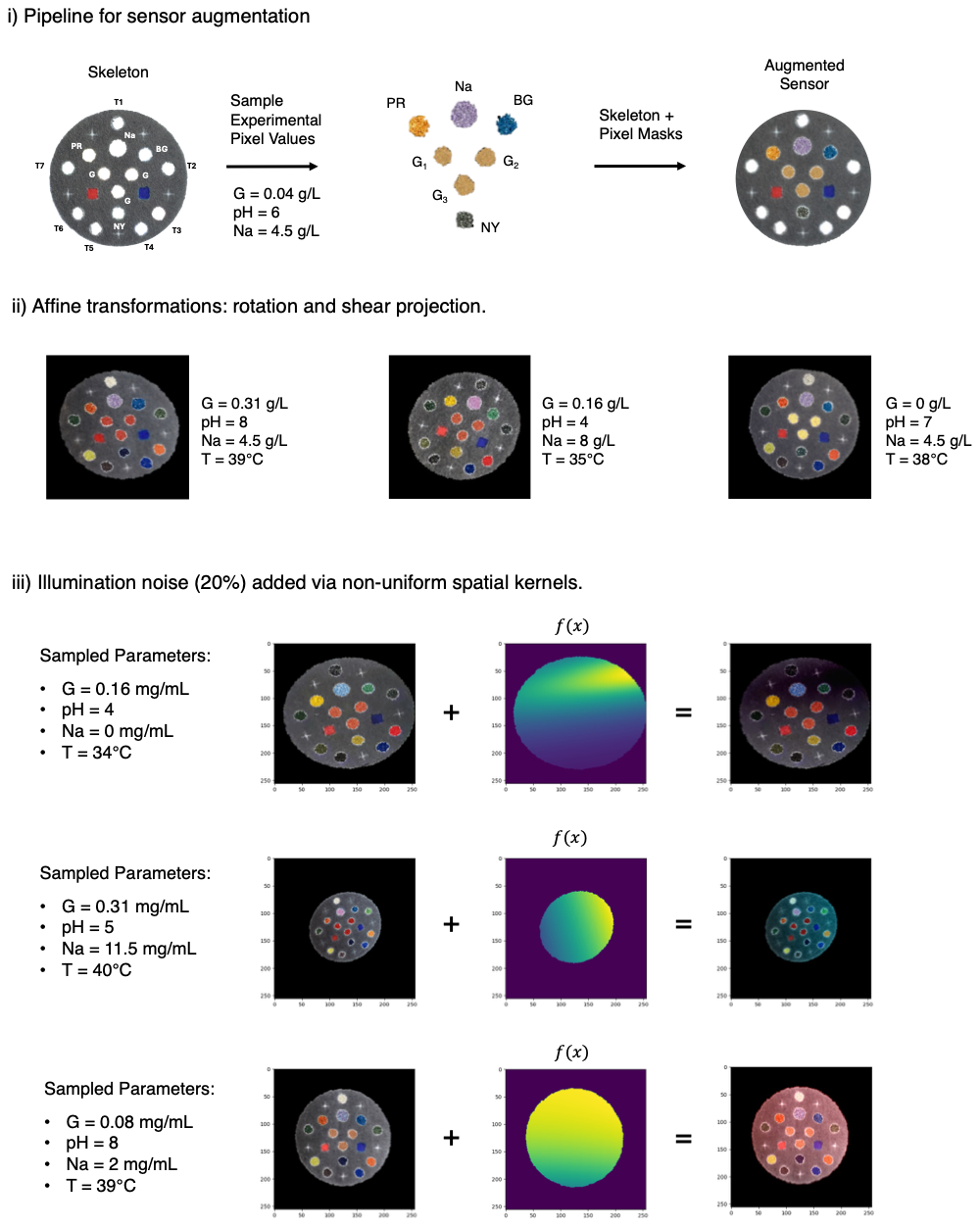

**C.** Illumination noise added via non-uniform spatial kernels.

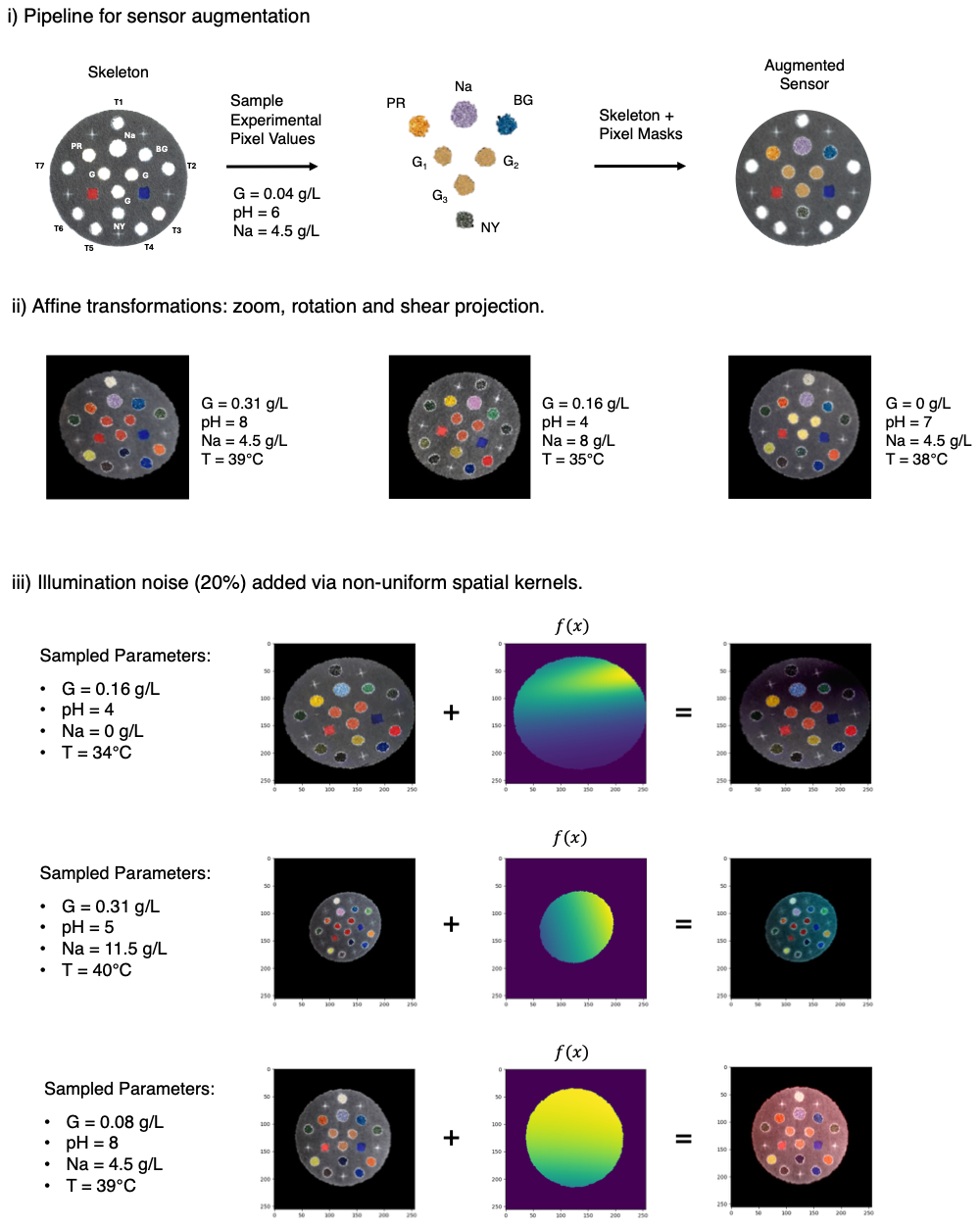

**Supplementary Figure 4**

**A.** Glucose Dye Colorimetric Distributions

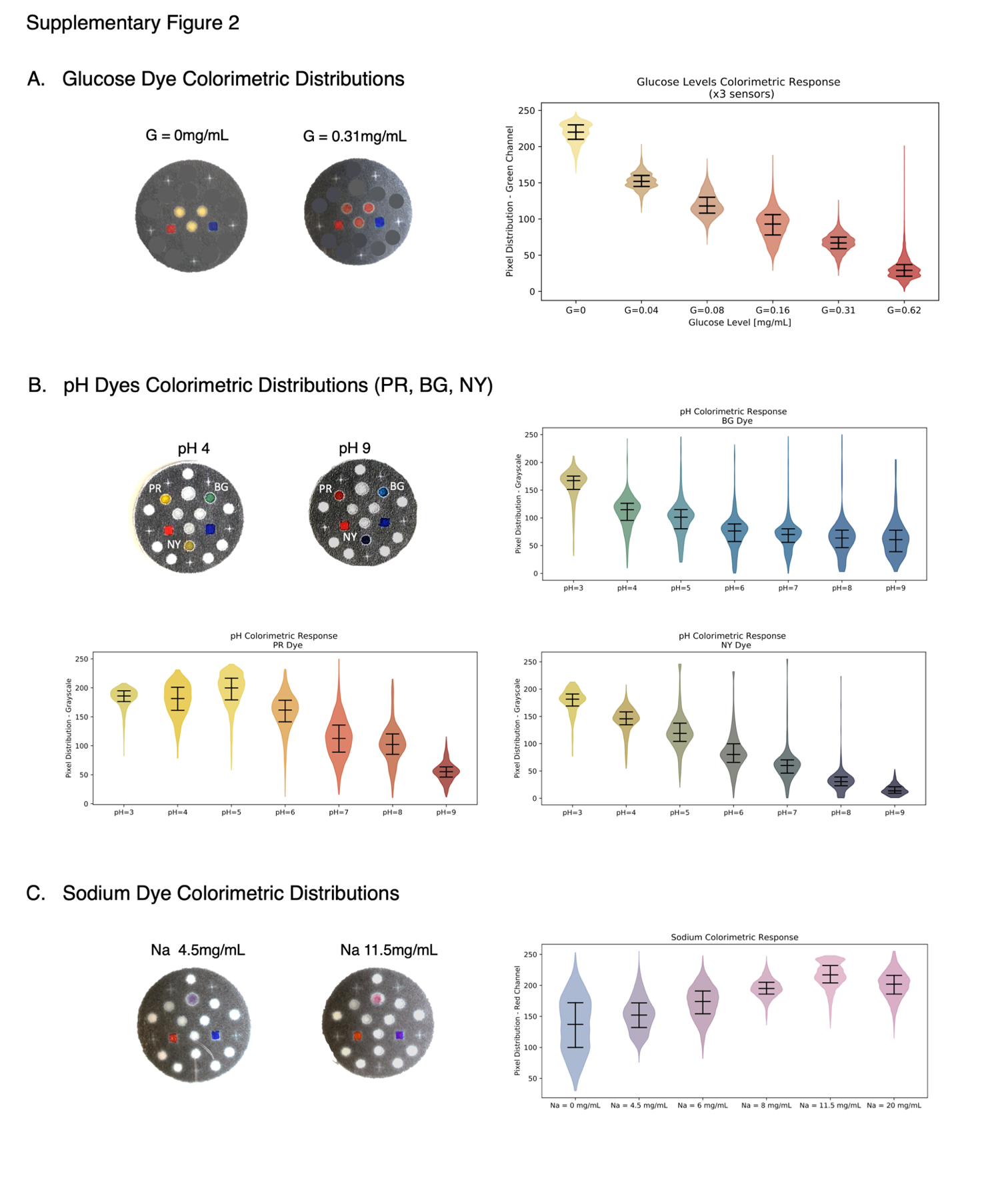

**B.** pH Dye Colorimetric Distributions (PR,BG,NY)

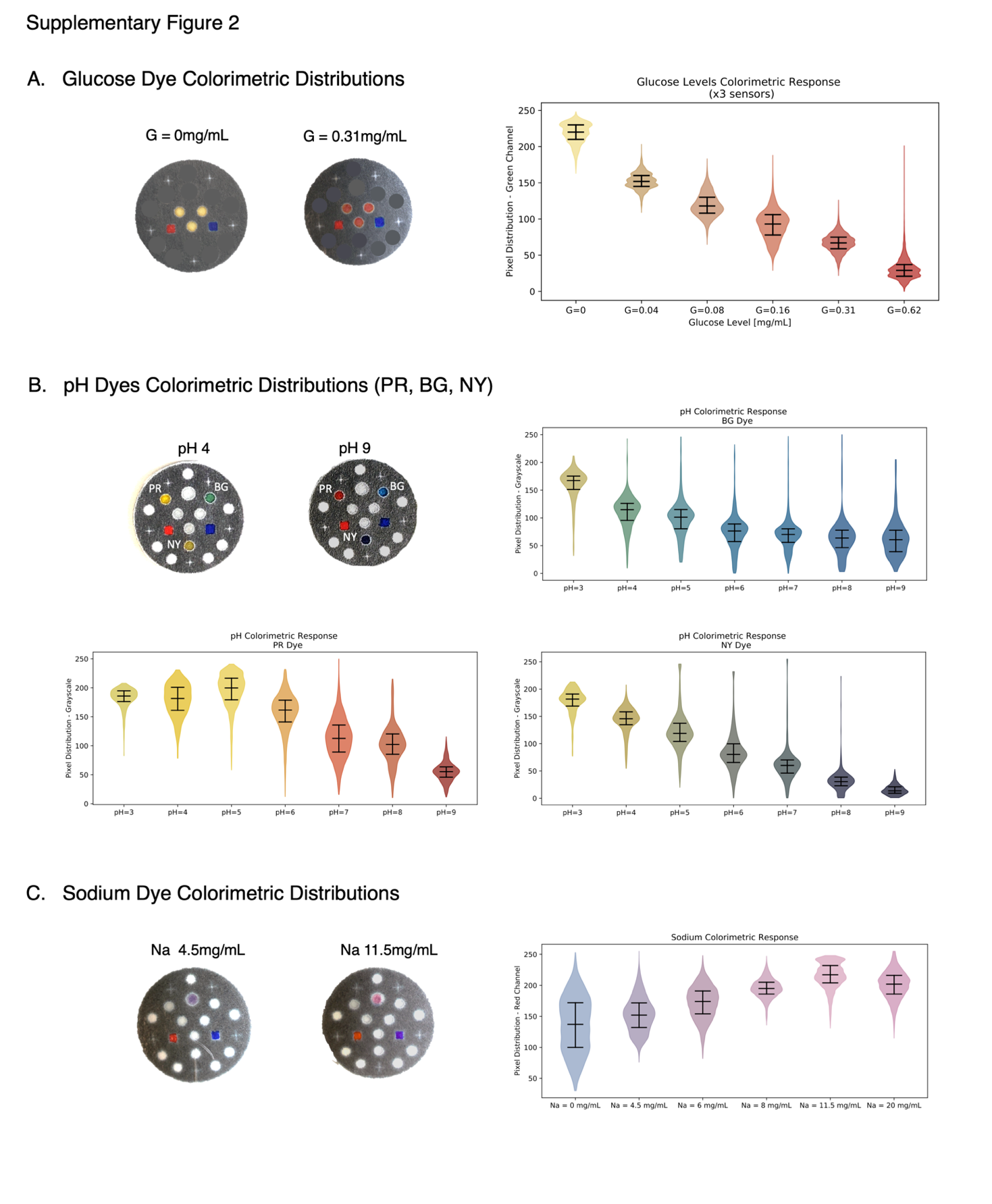

**C.** Sodium Dye Colorimetric Distributions

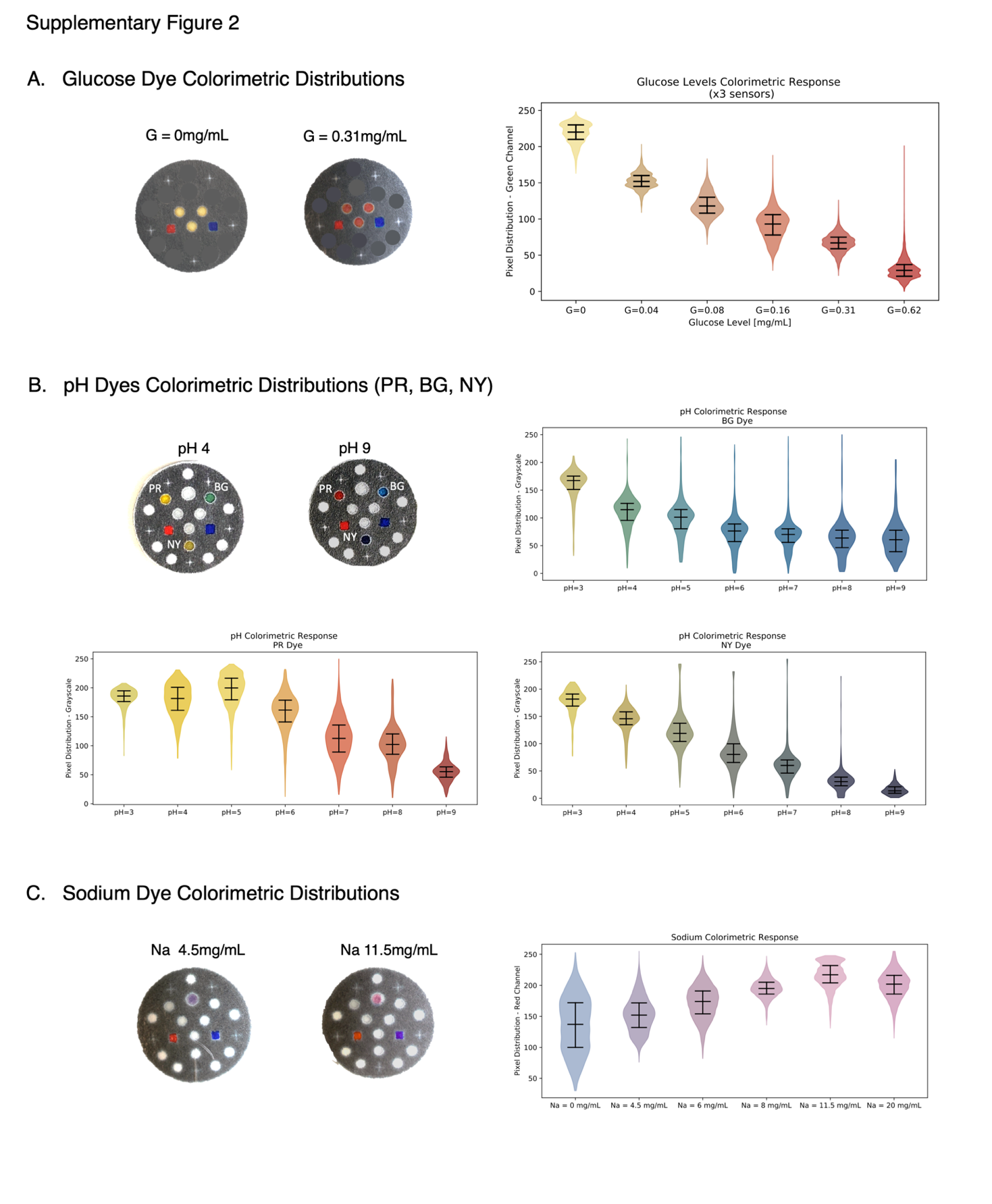

**Supplementary Figure 5**

**A.** Temperature Dyes (T1-T7) Colorimetric Distributions

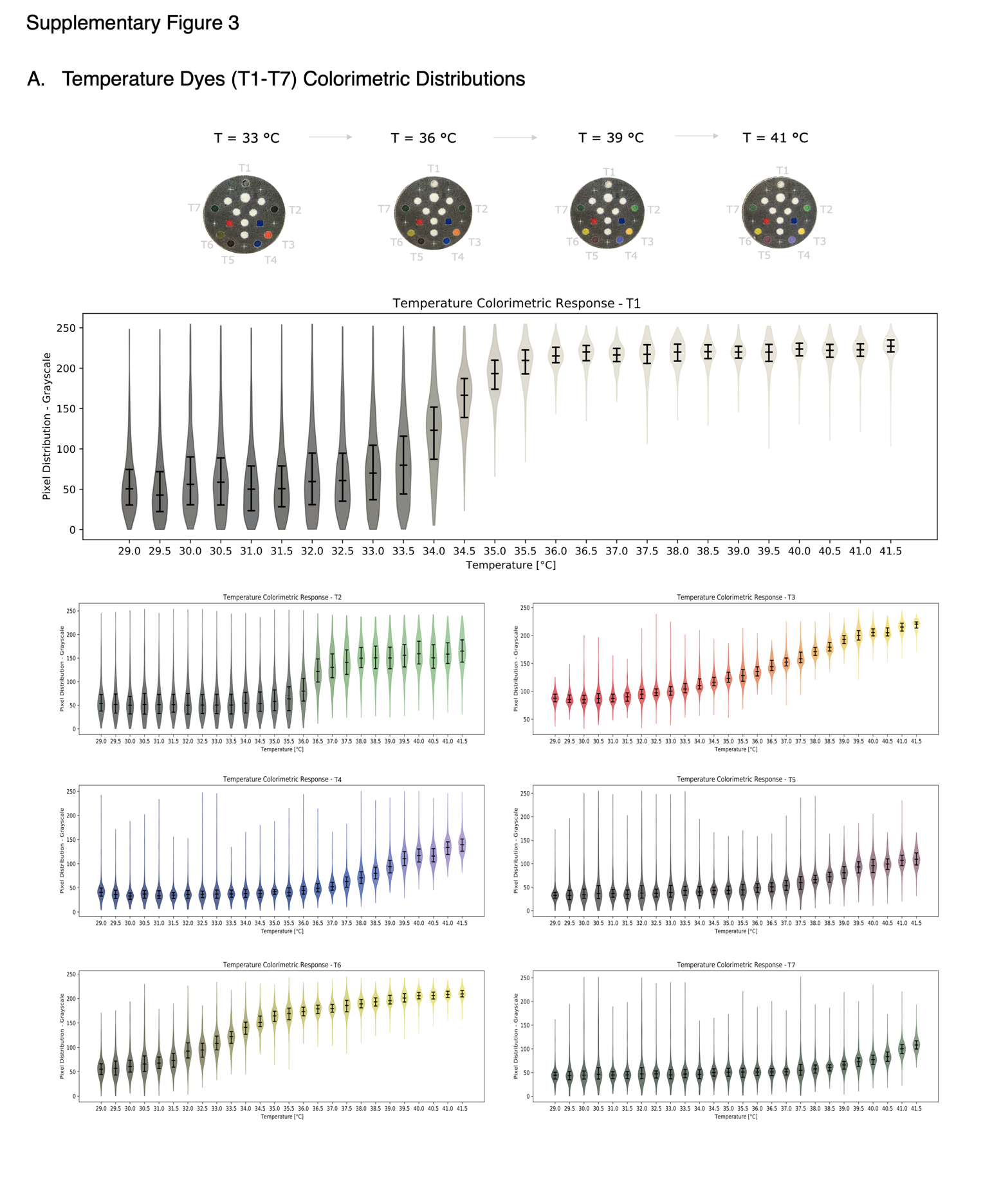

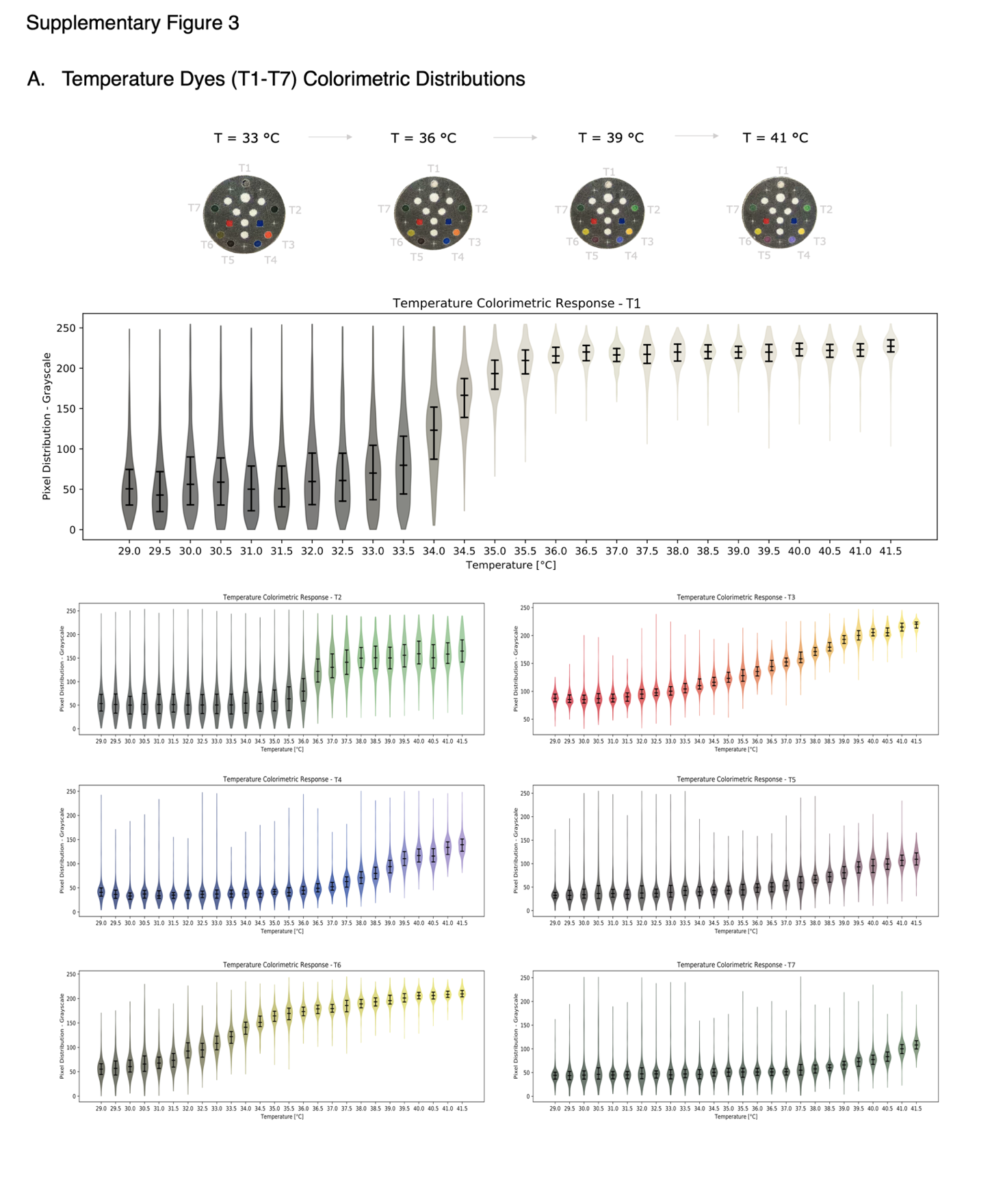

**Supplementary Figure 6**

**A.** Deep Learning Pipeline for Processing Experiment Videos

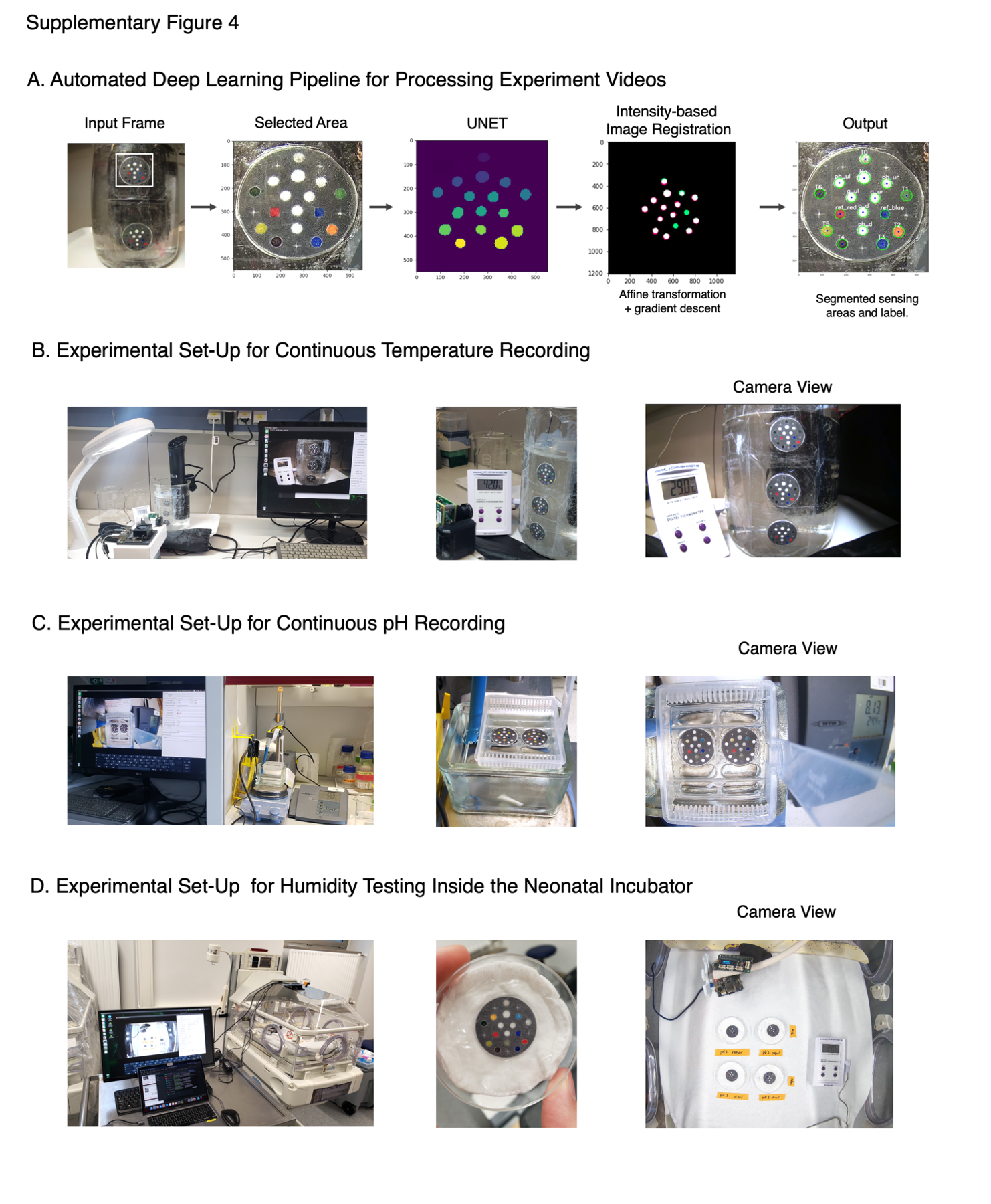

**B.** Experimental Set-Up for Continuous Temperature Recording

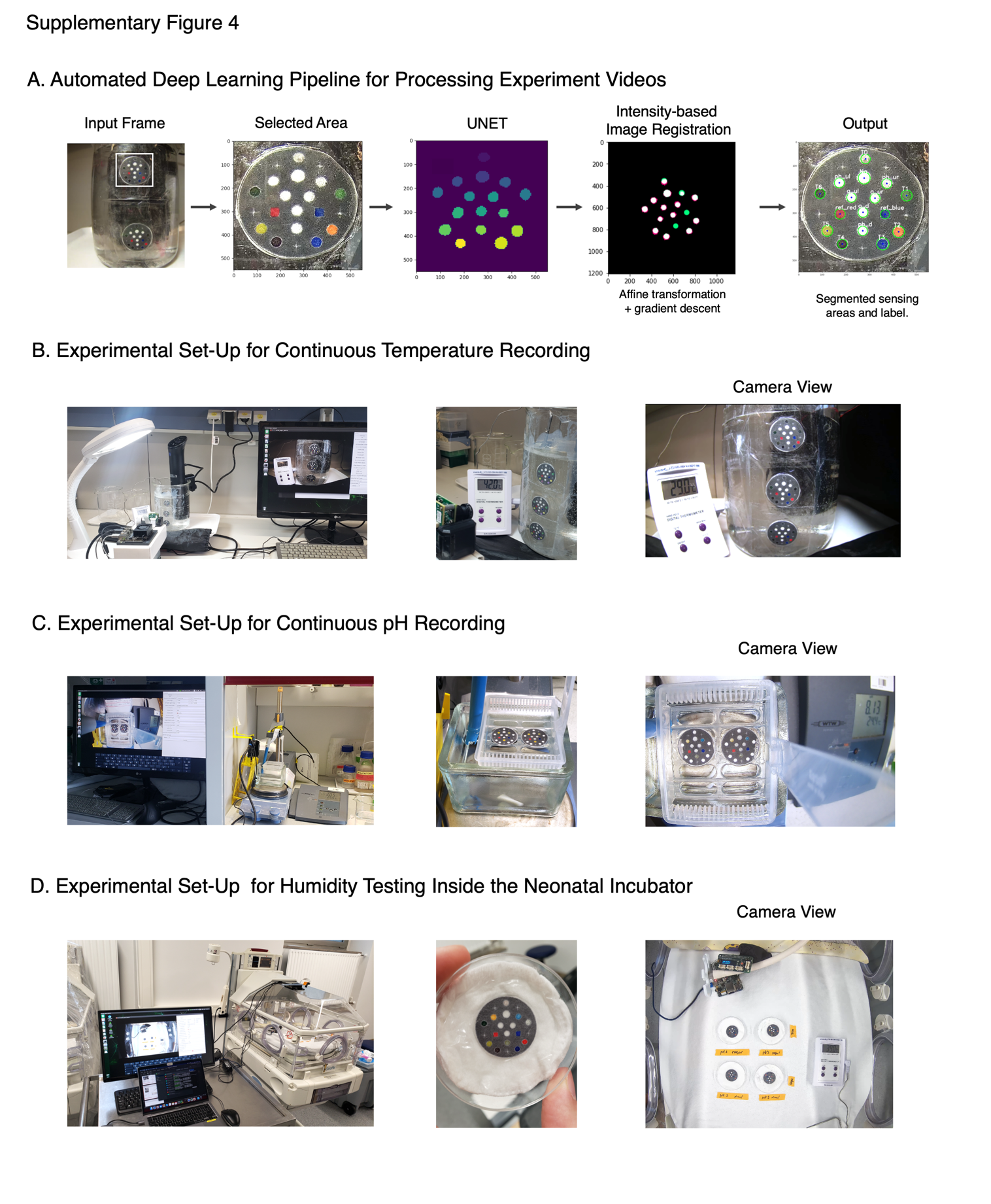

**C.** Experimental Set-Up for Continuous pH Recording

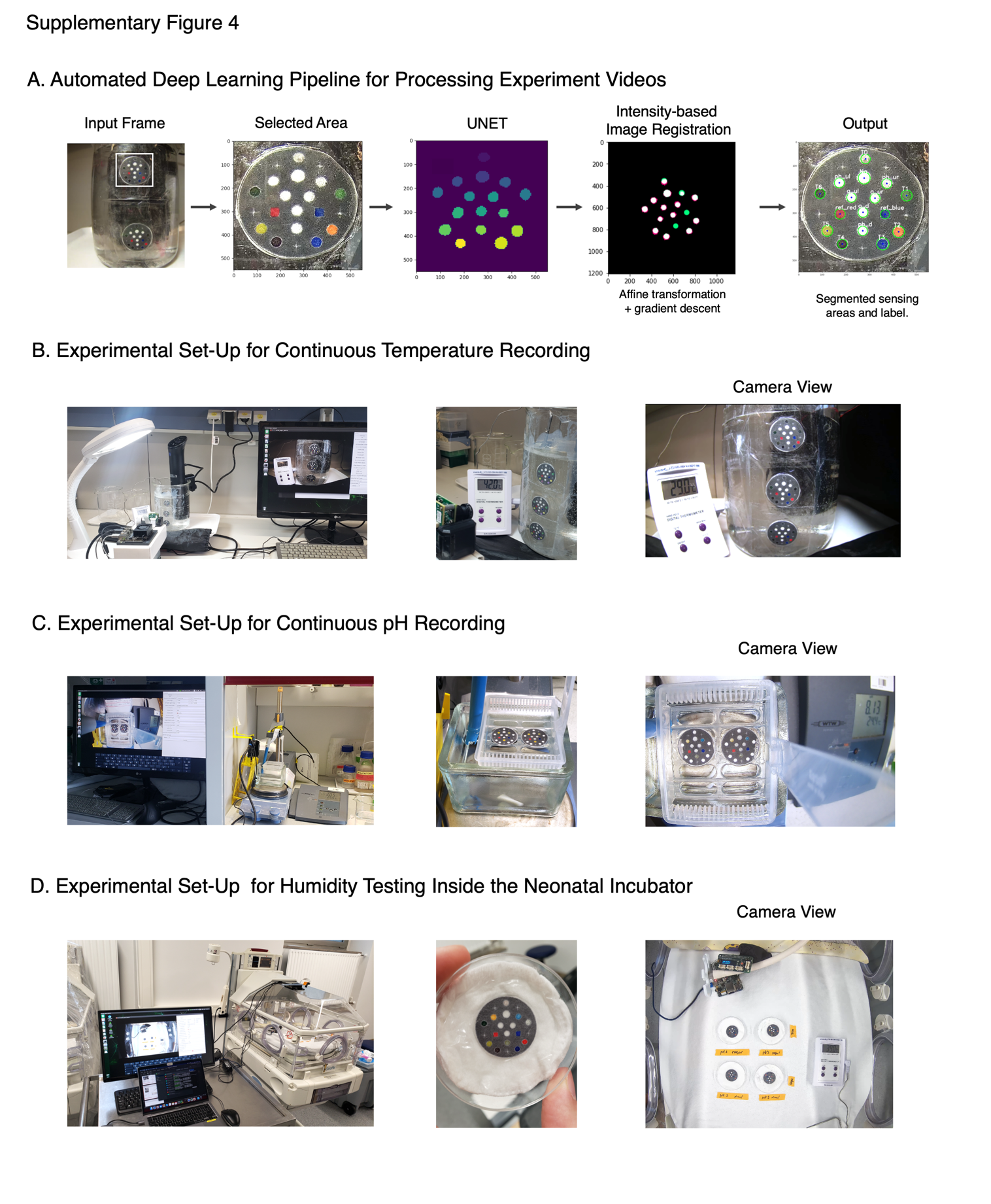

**D.** Experimental Set-Up for Humidity Testing Inside the Neonatal Incubator

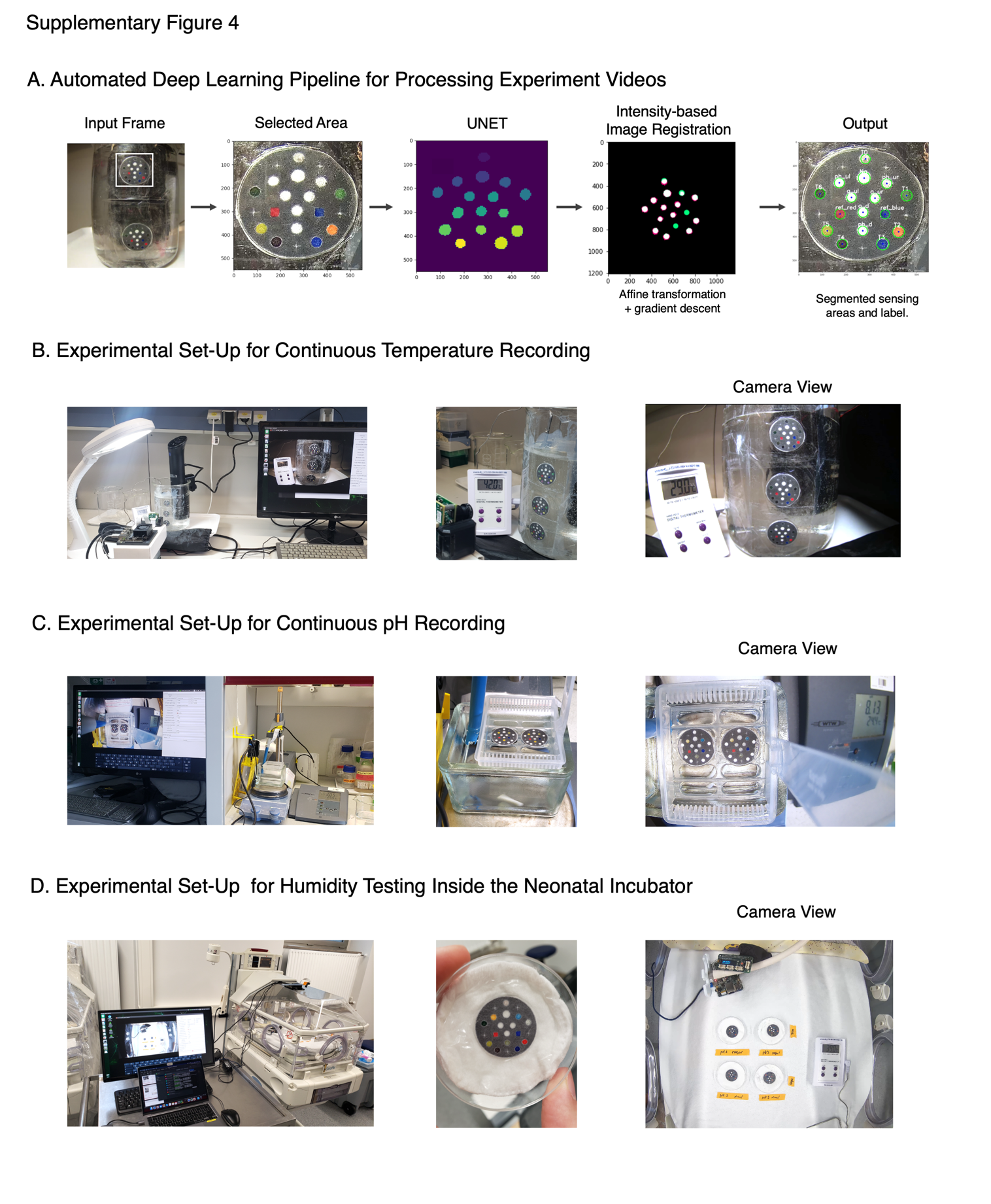

**Supplementary Figure 7**

**A.** Experimental Bench for Recording Inside the Neonatal Incubator

*i)* *Camera and Board Set-Up	ii) Baby breathing simulation*

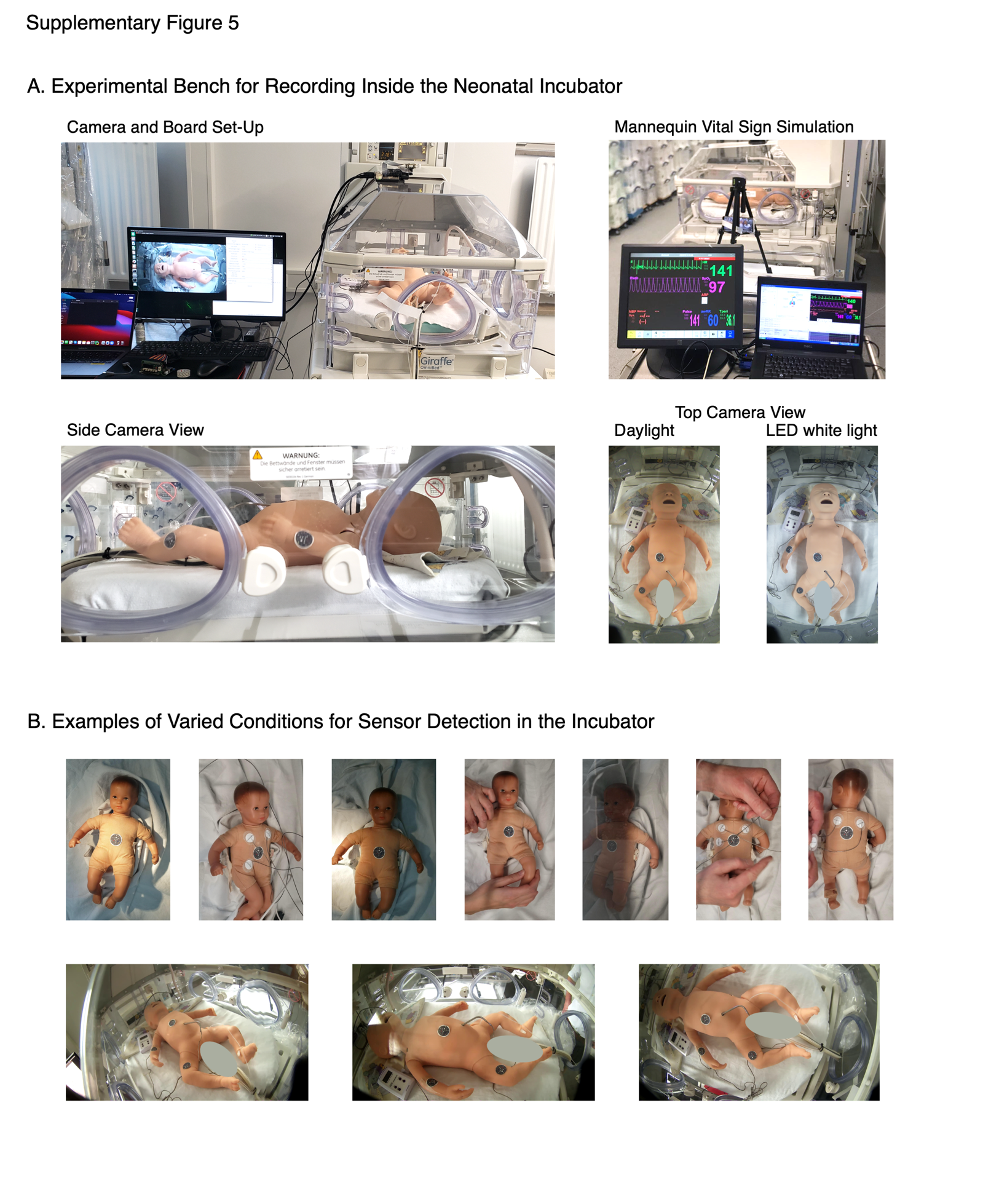

*i)* *Side Camera View	ii) Daylight (left), LED (right)*

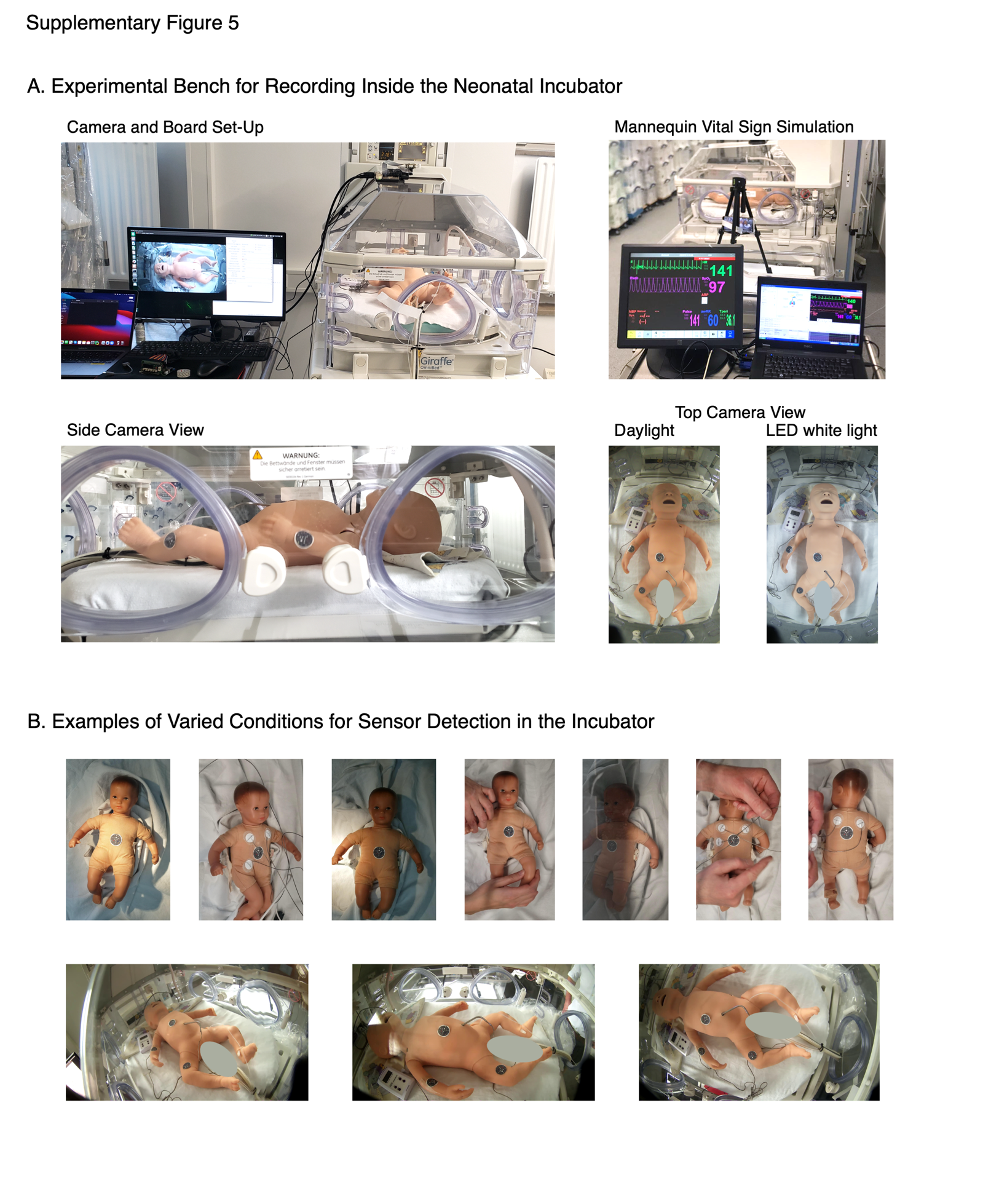

**B.** Examples of Varied Mannequin Conditions for Sensor Detection in the Incubator

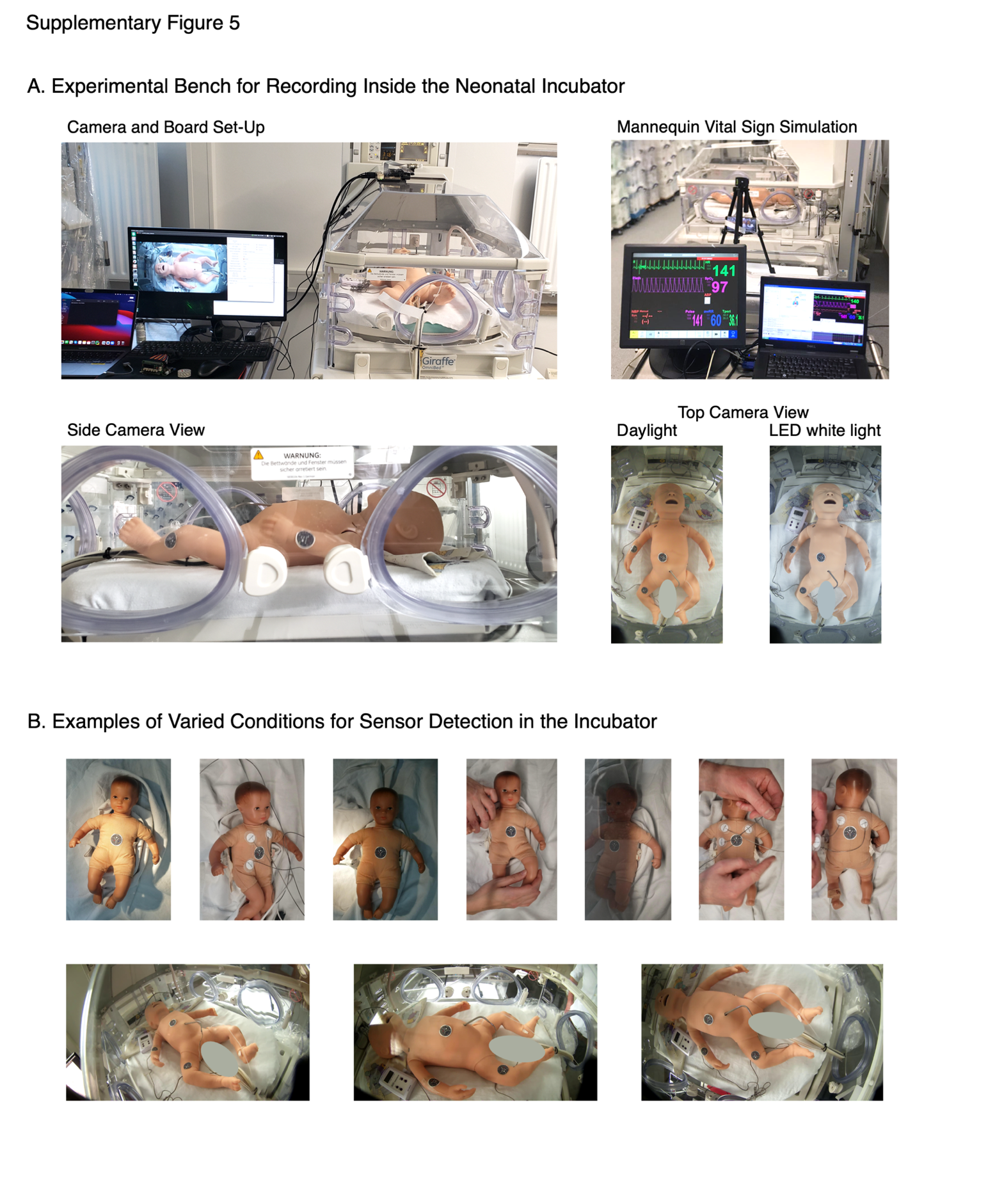
